# Critical assessment of the impact of vaccines and waning/boosting of immunity on the burden of COVID-19 in the U.S

**DOI:** 10.1101/2022.07.06.22277303

**Authors:** Calistus N. Ngonghala, Michael Asare-Baah

## Abstract

The COVID-19 pandemic continues to have a devastating impact on health systems and economies across the globe. Implementing public health measures in tandem with effective vaccination strategies have been instrumental in curtailing the burden of the pandemic. With the three vaccines authorized for use in the U.S. having varying efficacies and waning effects against major COVID-19 strains, understanding the impact of these vaccines on COVID-19 incidence and fatalities is critical. Here, we formulate and use mathematical models to assess the impact of vaccine type, vaccination and booster uptake, and waning of natural and vaccine-induced immunity on the incidence and fatalities of COVID-19 and to predict future trends of the disease in the U.S. when existing control measures are reinforced or relaxed. Results of the study show a 5, 1.8, and 2 times reduction in the reproduction number during the period in which vaccination, first booster, and second booster uptake started, respectively, compared to the previous period. Due to waning of vaccine-induced immunity, vaccinating up to 96% of the U.S. population might be required to attain herd immunity, if booster uptake is low. Additionally, vaccinating and boosting more people from the onset of vaccination and booster uptake, especially with mRNA vaccines (which confer superior protection than the Johnson & Johnson vaccine) would have led to a significant reduction in COVID-19 cases and deaths in the U.S. Furthermore, adopting natural immunity-boosting measures is important in fighting COVID-19 and transmission rate reduction measures such as mask-use are critical in combating COVID-19. The emergence of a more transmissible COVID-19 variant, or early relaxation of existing control measures can lead to a more devastating wave, especially if transmission rate reduction measures and vaccination are relaxed simultaneously, while chances of containing the pandemic are enhanced if both vaccination and transmission rate reduction measures are reinforced simultaneously. We conclude that maintaining or improving existing control measures and boosting with mRNA vaccines are critical in curtailing the burden of the pandemic in the U.S.

## 1. Introduction

The 2019 coronavirus (COVID-19) caused by severe acute respiratory syndrome coronavirus-2 (SARS-CoV-2) has been a significant global public health concern since it was first reported in December 2019 [1]. The World Health Organization (WHO) declared the outbreak a global pandemic on March 11, 2020 [2]. Although the implementation of non-pharmaceutical interventions (NPIs) and the roll-out of highly effective vaccines have been the mainstay public health strategy in curtailing the burden of the pandemic across the globe, the spread of the disease continues to have a devastating impact on healthcare systems and economies worldwide [3, 4], with 646, 637, 955 cases and 6, 645, 475 deaths as of early December 2022 [5]. The pandemic has had a ravaging effect in the United States (U.S.), the most impacted nation, with approximately 99, 087, 021 confirmed cases and 1, 082, 294 reported deaths as of early December 2022. [5, 6].

Vaccination stands out as a time-tested, cost-effective, and successful measure for curtailing the transmission of many infectious disease pathogens [7–9, 11]. Mass vaccination is one of the promising mitigating measures for COVID-19 and a strategy for generating herd immunity [12]. Currently, in the U.S., Pfizer-BioNTech, Moderna, and Janssen or Johnson & Johnson (J & J) vaccines have been approved by the Food and Drug Administration (FDA) for use against COVID-19 following multiple successful clinical trials [4, 16]. The FDA granted an emergency use authorization for Pfizer-BioNTech, Moderna, and Janssen vaccines on December 11, 2020, December 18, 2020, and February 27, 2021, respectively, [4], and final approval on August 23, 2021, for the Pfizer-BioNTech (COMIRNATY) vaccine, January 31, 2022, for the Moderna (Spikevax) vaccine [18] and limited use approval for the Janssen vaccine on May 5, 2022. The adenovirus-based Janssen vaccine requires a single dose, while the messenger ribonucleic acid (mRNA) Pfizer-BioNTech and Moderna vaccines require two doses to achieve full and durable protection. The initial dose is expected to prime the immune system, while the second dose provides an immune booster and enhances cross-protective activity against some variants of concern (VOC), like the Delta and Omicron variants. [20–22]. With high efficacies of ≈ 95% for the mRNA vaccines and ≈ 70% for the adenovirus-based vaccine against the non-variant (wild-type) strain of SARS-CoV-2, these vaccines have effectively prevented moderate to severe symptomatic disease, hospitalization, and death [11, 16, 20, 28, 29]. Although these vaccines were expected to contribute to halting the pandemic by driving the population to attain herd immunity [6, 30], their effectiveness in achieving this has been challenged by many factors, including the waning effects of the vaccines against major COVID-19 strains [3, 48], vaccine hesitancy [13, 15], and the emergence of new and resistant SARS-CoV-2 variants of concern that are more transmissible and to which existing vaccines do not offer sufficient cross-protection [3, 7, 12]. As of early December 2022, about 79% of the total U.S. population had received at least one dose of the recommended vaccines, with 68% fully vaccinated, and 33% having received at least one booster dose [19, 31].

Although the COVID-19 vaccines approved for use in the U.S. are effective against symptomatic laboratory-confirmed COVID-19 cases caused by the non-variant (wild-type) strain and some strains like the B.1.1.7 (Alpha), their efficacy may wane over time [3]. These vaccines have reduced potency against mutant variants like B.1.617.2 (Delta) and B.1.1.529 (Omicron), which are more virulent and highly transmissible and were widely in circulation in the U.S. at the time of this writing [30, 32]. Hence necessitating the administration of booster doses for conferring full or at least partial protection and mitigating the waning effect of the vaccine over time [25, 33]. Based on the evidence of breakthrough infections, the FDA, on September 22, 2021, authorized the use of booster doses for the Pfizer-BioNTech COVID-19 vaccine and subsequently issued approval for the Moderna and Janssen vaccines on October 20, 2021 [16, 65]. The first booster dose is administered as a single dose at least five months after completing the initial COVID-19 vaccination series of two doses for the mRNA vaccines and at least two months after the standard single dose for the Janssen vaccine. The CDC also recommends a second booster dose (fourth dose for mRNA vaccine receivers and third dose for Janssen vaccine receivers) for moderate or severely immunocompromised individuals 12 years and older. The second booster is recommended to be administered at least 3 months after the first booster shot. Except in some specific cases, mRNA vaccines must be used for the second booster [17]. To address the reduced efficacy of the monovalent vaccines against SARS-CoV-2 variants of concern, such as the Omicron variant, the Advisory Committee on Immunization Practices recommended the use of bivalent Pfizer-BioNTech and Moderna (mRNA) vaccines on September 1, 2022. Compared with the monovalent mRNA vaccines, the bivalent mRNA booster has extended immunity and provides additional protection against symptomatic SARS-CoV-2 infection for individuals who had previously received 2, 3, or 4 monovalent vaccine doses [57]. These bivalent mRNA vaccines are effective against the wild-type SARS-CoV-2 strain and the BA.4 and BA.5 Omicron sub-variants. Until October 12, 2022, when these bivalent mRNA vaccines were recommended for boosting younger individuals (between the ages of 5 and 11 years old), only individuals aged 12 and above (for the bivalent Pfizer-BioNTech vaccine) and 18 years and above (for the Moderna vaccine), who had completed the primary series of any of the three monovalent vaccines were eligible for the bivalent vaccines approved for use against COVID-19 in the U.S. [14, 57].

Several mathematical modeling frameworks have been used to understand the impact of vaccination on the transmission dynamics of COVID-19. A two-strain and two-group mathematical model was developed in [34] and used to assess the impact of vaccination and vaccine-induced cross-protection against the B.1.1.7 and other SARS-CoV-2 variants circulating in the U.S. The study shows that future waves of the COVID-19 pandemic can be prevented in the U.S. if the existing vaccines offer a moderate level of cross-protection against the variant. Iboi *et al*. [35] used a deterministic model to assess the impact of a hypothetical imperfect COVID-19 vaccine on the transmission dynamics of the COVID-19 pandemic in the U.S. Their results show that the prospect of eliminating the local transmission of COVID-19 in the U.S. using the hypothetical vaccine is greatly enhanced if the vaccination program is combined with other interventions such as face mask usage and/or social distancing. A compartmental model was developed in [36] and associated with COVID-19 data from Italy to compute the time profile of healthcare system costs, hospitalization, and intensive care unit occupancy and deaths. The model was also used to compare different vaccination scenarios and to assess the effect of mass vaccination campaigns as a function of the reproduction number due to SARS-CoV-2 variants. A two-group mathematical model (based on face-mask use in public) for assessing the population-level impact of the approved COVID-19 vaccines on the COVID-19 pandemic was developed and analyzed in [38]. The study shows that the waning of natural and vaccine-induced immunity against COVID-19 induces only a marginal increase in the burden and the time-to-elimination of the pandemic. Moore *et al*. [39] used an age-structured vaccination model to assess the possibility of SARS-CoV-2 mortality or quality-adjusted life-year (QALY) losses in the UK. Their results show that vaccinating the older population has the most significant impact in reducing mortality. Islam *et al*. [40] used a model that accounted for the influence of age stratification and time-dependent infectivity to evaluate various vaccination strategies in the U.S. Their findings suggest that the CDC’s vaccine-allocation strategy is not optimal. Ngonghala *et al*. [25] developed and used a two-group model to explore the dynamics of two co-circulating variants (Delta and Omicron) of the SARS-CoV-2 virus in the presence of vaccination, waning vaccine-induced immunity, masking, and anti-viral treatment in the U.S. They showed that vaccination, combined with surgical or N95 masks increases the likelihood of containing COVID-19 in the U.S. and that although N95 masks are more efficient than surgical masks, more surgical mask-use is effective in combating COVID-19 than less N95 mask-use. Taboe *et al*. developed and used a mathematical framework structured by age and vaccination status to assess the impact of age structure and vaccine prioritization on COVID-19 in West Africa. Their findings suggested that age structure is critical in explaining the low incidence of COVID-19 in West Africa and that compliance with vaccination and NPI use is imperative for containing the pandemic in the region [24]. Other studies, including those in [10, 47, 49] have developed and used mathematical models structured by age or risk of contracting/spreading COVID-19 to assess the impact of booster vaccination strategies. These studies highlight the importance of prioritizing vaccination and effective implementation of NPIs as critical for containing the pandemic. However, none of these studies investigated the impact of using specific vaccines (e.g., the J & J versus mRNA vaccines) used for vaccination and boosting explicitly or the impact of vaccine/booster timing on the dynamics of COVID-19.

In this study, we develop and use a model framework (that is structured by vaccine type) with data on confirmed new daily and cumulative COVID-19 cases and deaths for the U.S. to assess the impact of 1) full vaccination with the one-dose J & J and the two-dose mRNA (Pfizer-BioNTech and Moderna) vaccines licensed for use in the U.S.; 2) first and second booster uptake with the one-dose J & J or the two-dose mRNA vaccines; 3) early implementation of vaccine and boosting measures; 4) relaxation or reinforcement of vaccine and transmission rate reduction measures such as masking-up; and 5) waning natural and vaccine-induced immunity on the burden and future trajectory of COVID-19 in the U.S. (driven by the major circulating variants). The models are developed in Section 2. Analytical and numerical simulation results are presented in Section 3, and a discussion, limitations, and concluding remarks are presented in Section 4.

## 2. Methods

### 2.1. Model formulation

We develop four models to assess the dynamics of COVID-19 in the U.S.: 1) a basic model with no vaccination, 2) an addon to the basic model that accounts for one- and two-dose vaccines and the waning effect of vaccine-induced immunity, 3) the model in 2) that accounts for one booster vaccine dose, and 4) the model in 3) with a second booster vaccine dose.

### 2.2. Model 1: The basic model

The basic model is the typical transmission model of the virus in a human population, including a confirmed case class. Here, the total human population (*N*) is broken down into susceptible (*S*), exposed (*E*), pre-symptomatic infectious (*I*_*p*_), Symptomatic infectious (*I*_*s*_), asymptomatic infectious (*I*_*a*_), confirmed (*I*_*c*_), hospitalized (*I*_*h*_), and recovered (*R*) individuals. All human recruitments are into the susceptible class at a rate Λ *humans per day*, while natural death in each of the classes is at per capita rate *μ per day* (i.e., 1/*μ* is the average life span of an individual). Susceptible individuals acquire the infection through contacts with *I*_*p*_, *I*_*s*_, *I*_*a*_, *I*_*c*_, *I*_*h*_ individuals at per capita rate *β*_*p*_ *I*_*p*_ /*N, β*_*s*_ *I*_*s*_ /*N, β*_*a*_ *I*_*a*_ /*N, β*_*c*_ *I*_*c*_ /*N*, and *β*_*h*_ *I*_*h*_ /*N per day*, respectively. Hence, the rate at which susceptible individuals are infected (i.e., the force of infection) is:

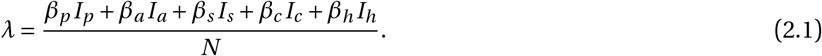

Although confirmed and hospitalized individuals are expected to be isolated from the actively-mixing population, in reality, isolation mandates are not always perfectly respected. This gives rise to the concept of “leaky isolation”, in which some confirmed cases (especially those who are not hospitalized), who are supposed to be isolated, are not completely isolated for various reasons, including economic hardship and human behavioral change. In the current framework, we factor in the possibility that not all confirmed and hospitalized individuals comply with isolation mandates strictly. This allows for the possibility of some level of disease transmission by confirmed and hospitalized individuals. Accordingly, the estimated transmission rates for confirmed and hospitalized individuals are much smaller than those for unconfirmed infectious individuals, who are not supposed to be isolated (see Tables S4-S8 of the SI). Exposed individuals progress to the pre-symptomatic infectious class at per capita rate *σ*_*e*_ *per day*, (i.e., 1/*σ*_*e*_ is the average latent period), while pre-symptomatic infectious individuals progress to the symptomatic infectious class at rate (1 − *r*)*σ*_*p*_ *per day* or to the asymptomatic infectious class at per capita rate *rσ*_*p*_ *per day*, where 0 < *r* ≤ 1 (0 < 1 − *r* ≤ 1) is the proportion of pre-symptomatic infectious individuals who develop (do not develop) disease symptoms at the end of the incubation period and 1/*σ*_*p*_ is the average pre-symptomatic infectious period. The compartment for confirmed cases (*I*_*c*_) consists of confirmed COVID-19 cases from prevailing public health data and therefore is important for fitting our model to the observed data. This class is populated by individuals, who test positive for COVID-19 from the *I*_*p*_, *I*_*a*_, or *I*_*s*_ class at per capita rate *τ*_*p*_, *τ*_*a*_, or *τ*_*s*_ *per day*, respectively. Individuals from this class either become hospitalized (or are treated in a healthcare setting) at per capita rate *ρ*_*c*_ *per day* (i.e., 1/*ρ*_*c*_ is the average length of time that elapses before confirmed individuals are hospitalized or seek treatment), or recover from infection to join the recovered class (*R*) at per capita rate, *γ*_*c*_ *per day* (1/*γ*_*c*_ is the average duration of the infectious period for confirmed cases). Symptomatic (asymptomatic) infectious individuals recover at per capita rate *γ*_*s*_ (*γ*_*a*_) *per day*, while natural immunity wanes at per capita rate *ω*_*r*_ *per day* (i.e., 1/*ω*_*r*_ is the average duration of natural immunity to COVID-19). Individuals in the *I*_*k*_, *k* ∈ {*c, h, s*} class die from COVID-19 at per capita rate *δ*_*k*_ *per day*. The flow diagram of the model is presented in Fig. 1, while the variables and parameters are further described in Tables S1 and S2 of the online supplementary information (SI). The dynamics of the susceptible (*S*) and exposed (*E*) populations are described by the equations:

**Fig. 1:**
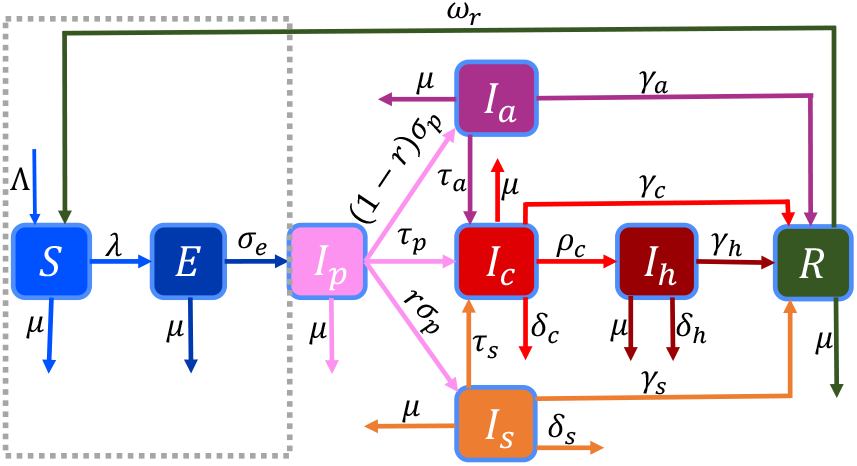
Schematic depiction of the basic model. The population is partitioned into susceptible (*S*), exposed (*E*), pre-symptomatic infectious (*I*_*p*_), Symptomatic infectious (*I*_*s*_), asymptomatic infectious (*I*_*a*_), confirmed (*I*_*c*_), hospitalized (*I*_*h*_), and recovered (*R*) individuals. The model parameters are described in the text and Table S2 of the SI.

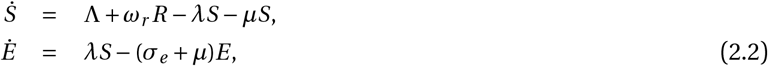

while the dynamics of the presymptomatic infectious (*I*_*p*_), asymptomatic infectious (*I*_*a*_), symptomatic infectious (*I*_*s*_), confirmed cases (*I*_*c*_), hospitalized (*I*_*h*_), and recovered (*R*), are described by the equations:

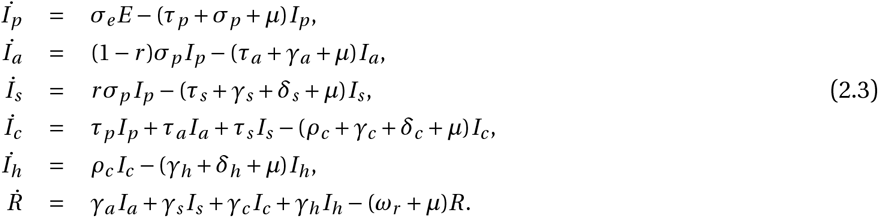

Hence, the basic model (Model 1) is governed by Eqs. (2.2)-(2.3). Since Eqs. (2.3) (discussed in this section) are common to all the models formulated in this study; we will focus subsequent model description on the additional classes generated from the susceptible class as a result of vaccination and terms from these classes that feed into the latent class. That is, descriptions of subsequent models will focus on new classes within the region in the dotted box in Fig. 1.

### 2.3. Model 2: The vaccination model (i.e., the basic model with vaccination and waning vaccine-induced immunity)

In this section, the basic model (Eqs. (2.2)-(2.3)) is extended to account for vaccination of susceptible individuals using the three vaccines authorized for use in the U.S. Since the Pfizer-BioNTech and the Moderna vaccines are both administered in two-doses and have efficacies of 95% and 94.1%, respectively, we group the two vaccines and consider an average of the two efficacies (i.e., (0.95 + 0.941)/2 = 0.9455) for this class. Furthermore, because the duration of vaccine-induced immunity for these two vaccines is similar, it is reasonable to group individuals who receive them together. Hence, the extended model accounts for susceptible individuals vaccinated with either the J & J vaccine (administered as a single dose) and the Pfizer-BioNTech or Moderna vaccine (administered in two doses). The susceptible population (*S*, from the basic model) is split into individuals who are unvaccinated (*S*_*u*_), vaccinated with the J & J vaccine (*S*_*j*1_) at per capita rate (*ξ*_*ju*_ *per day*), vaccinated with the first dose of an mRNA vaccine, i.e., either the Pfizer-BioNTech or Moderna vaccine (*S*_*m*1_) at par capita rate (*ξ*_*mu*_ *per day*), and vaccinated with a second dose of the same mRNA vaccine as the first dose (*S*_*m*2_) at per capita rate (*ξ*_*m*1_ *per day*). It is assumed that individuals in the *S*_*j*1_ (*S*_*m*2_) class progress to a temporary class, *S*_*jw*1_ (*S*_*mw*1_) at per capita rate *α*_*j*1_ (*α*_*m*2_) *per day* when their vaccine-induced immunity starts waning. It should be mentioned that waning of vaccine-induced immunity is a continuous process and can be modeled as a dynamic variable that changes with time or using multiple waning classes. However, for parsimony and tractability, we use only one class in each waning scenario. Individuals from the *S*_*jw*1_ (*S*_*mw*1_) class progress to the *S*_*u*_ class at per capita rate, *α*_*jw*1_ (*α*_*mw*1_) *per day*, when their vaccine-induced immunity wanes completely, while individuals who received only a single dose of one of the mRNA vaccines progress to the *S*_*u*_ class at per capita rate (*α*_*m*1_), when their vaccine-induced immunity wanes completely. Break-through infections for individuals in the *S*_*l*_, *l* ∈ { *j* 1, *m*1, *m*2, *mw* 1, *jw* 1} class are at rate (1−*ε*_*l*_)*λ per day*, where *λ* is as defined in Eq. (2.1), and 0 ≥ *ε*_*l*_ ≥ 1 is the efficacy of vaccines in preventing infections. The other variables and parameters are as described for the basic model (Eqs. (2.2)-(2.3)), and the total population (*N*) is given by *N* = *S*_*u*_ + *S*_*j*1_ + *S*_*m*1_ + *S*_*m*2_ + *S*_*mw*1_ + *S*_*jw*1_ + *E* + *I*_*p*_ + *I*_*a*_ + *I*_*s*_ + *I*_*c*_ + *I*_*h*_ + *R*. Schematics of the unvaccinated susceptible, vaccinated, and exposed portion of the model are presented in Fig. 2; the corresponding equations are given in Eqs. (2.4), and the full vaccination model is described by Eqs. (2.2) and (2.4).

**Fig. 2:**
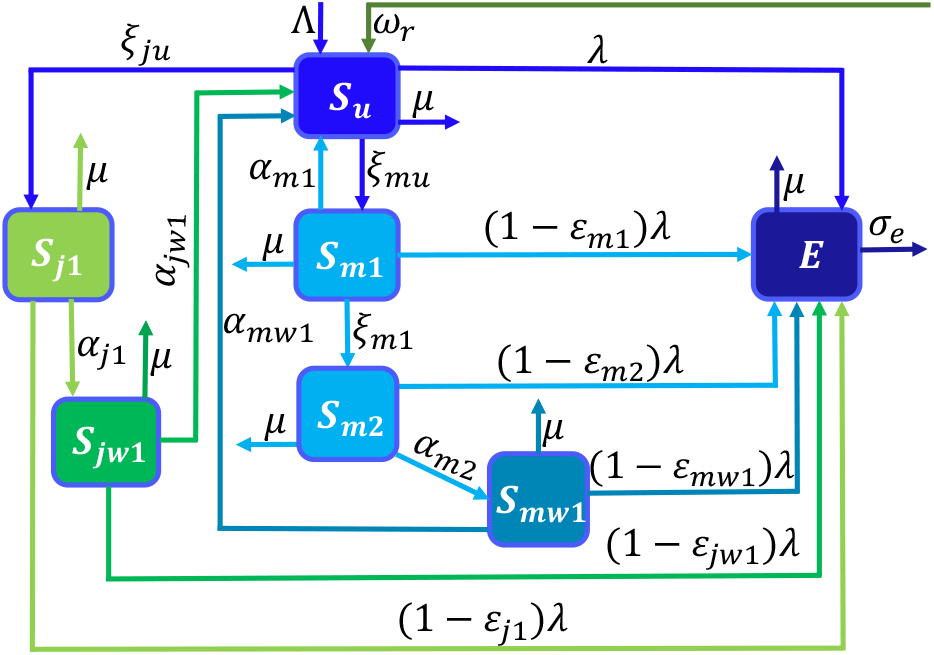
Schematics of the vaccination model, i.e., an extension of the basic model given by Eqs. (2.2)-(2.3) to include individuals who are not vaccinated (*S*_*u*_), vaccinated with the J & J vaccine (*S*_*j*1_) and individuals, *S*_*m*1_ (*S*_*m*2_), who have received the first (second) dose of an mRNA (Pfizer-BioNTech or Moderna) vaccine. Individuals who are fully vaccinated with the J & J (an mRNA) vaccine progress to the *S*_*jw*1_ (*S*_*mw*1_) class when their immunity wanes. The rest of the schematics, i.e., the portion from the exposed through the recovered class (not shown here), is as in Fig. 1. The variables and parameters are described in the text and in Tables S1-S2 in the SI.

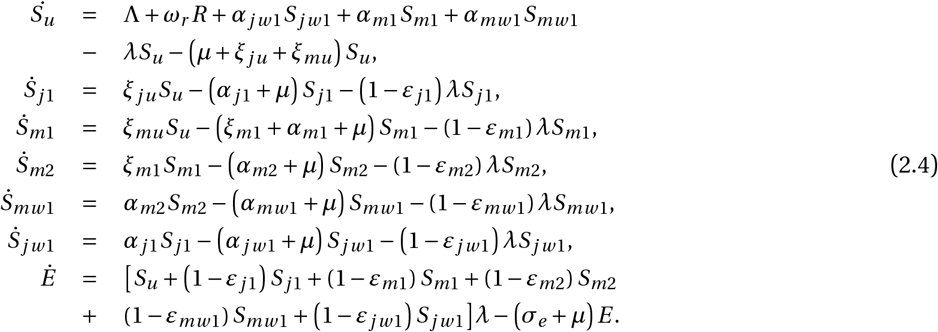

### 2.4. Model 3: The vaccination model (i.e., Model 2) with first booster vaccine dose

The vaccination model from Section 2.2 (i.e., Eqs. (2.2) and (2.4)) is extended to account for the first booster vaccine dose uptake for individuals who received the one dose J & J vaccine at least two months earlier (*S*_*jw*1_), or individuals who received a second dose of any of the mRNA vaccines at least five months earlier (*S*_*mw*1_). Individuals (*S*_*jw*1_) who received the J & J vaccine at least two months earlier are boosted with either the J & J vaccine (*S*_*j* 2_) at per capita rate (*ξ*_*jw*1_ *per day*), or with one of the mRNA vaccines (*S*_*jmw*_) at per capita rate (*ξ*_*jm*1_ *per day*), while individuals who were fully vaccinated with an mRNA vaccine, i.e., individuals who have received two doses of the same mRNA vaccine at least five months earlier (*S*_*mw*1_), are boosted with the same mRNA vaccine (*S*_*m*3_) at per capita rate (*ξ*_*mw*1_ *per day*). When vaccine-induced immunity of individuals, who were fully vaccinated and boosted with the J & J vaccine wanes, they progress to the *S*_*jw* 2_ class at per capita rate (*α*_*jw* 2_ *per day*), while individuals who were fully vaccinated with the J & J vaccine, but boosted with one of the mRNA vaccines progress to the *S*_*jmw*_ class at per capita rate (*α*_*jmw*_ *per day*) when their immunity start waning. When vaccine-induced immunity of individuals who were fully vaccinated and boosted with an mRNA vaccine wanes, the individuals progress to the *S*_*mw* 2_ class at per capita rate (*α*_*m*3_ *per day*). Individuals in the *S*_*jw* 2_ and *S*_*mw* 2_ classes progress to the *S*_*u*_ class at per capita rates *α*_*jw* 2_ and *α*_*jw* 2_, respectively, when their vaccine-induced immunity wanes completely. Break-through infections in these new classes are at rates (1 − *ε*_*l*_)*λ*, where *λ* is as defined in Eq. (2.1), and 0 ≥ *ε*_*l*_ ≥ 1, *l* ∈ { *j* 2, *jm*1, *jw* 2, *m*3, *mw* 2, *jmw* } are the efficacies of the booster vaccine doses in preventing infections. The other variables and parameters are as defined in Sections 2.2 and 2.3 and Tables S1-S2 of the SI, while the total population (denoted by *N*) is given by *N* = *S*_*u*_ + *S*_*j*1_ + *S*_*m*1_ + *S*_*m*2_ + *S*_*mw*1_ + *S*_*jw*1_ + *S*_*j* 2_ + *S*_*jm*1_ + *S*_*jw* 2_ + *S*_*m*3_ + *S*_*mw* 2_ + *S*_*jmw*_ + *E* + *I*_*p*_ + *I*_*a*_ + *I*_*s*_ + *I*_*c*_ + *I*_*h*_ + *R*. Schematics of the unvaccinated susceptible, vaccinated, boosted, and exposed portions of Model 3 (i.e., Model 2 with the first booster dose) are presented in Fig. 3. The model equations are given in Eqs. (2.5), and the full vaccination model with one booster dose uptake is described by Eqs. (2.2) and (2.5).

**Fig. 3:**
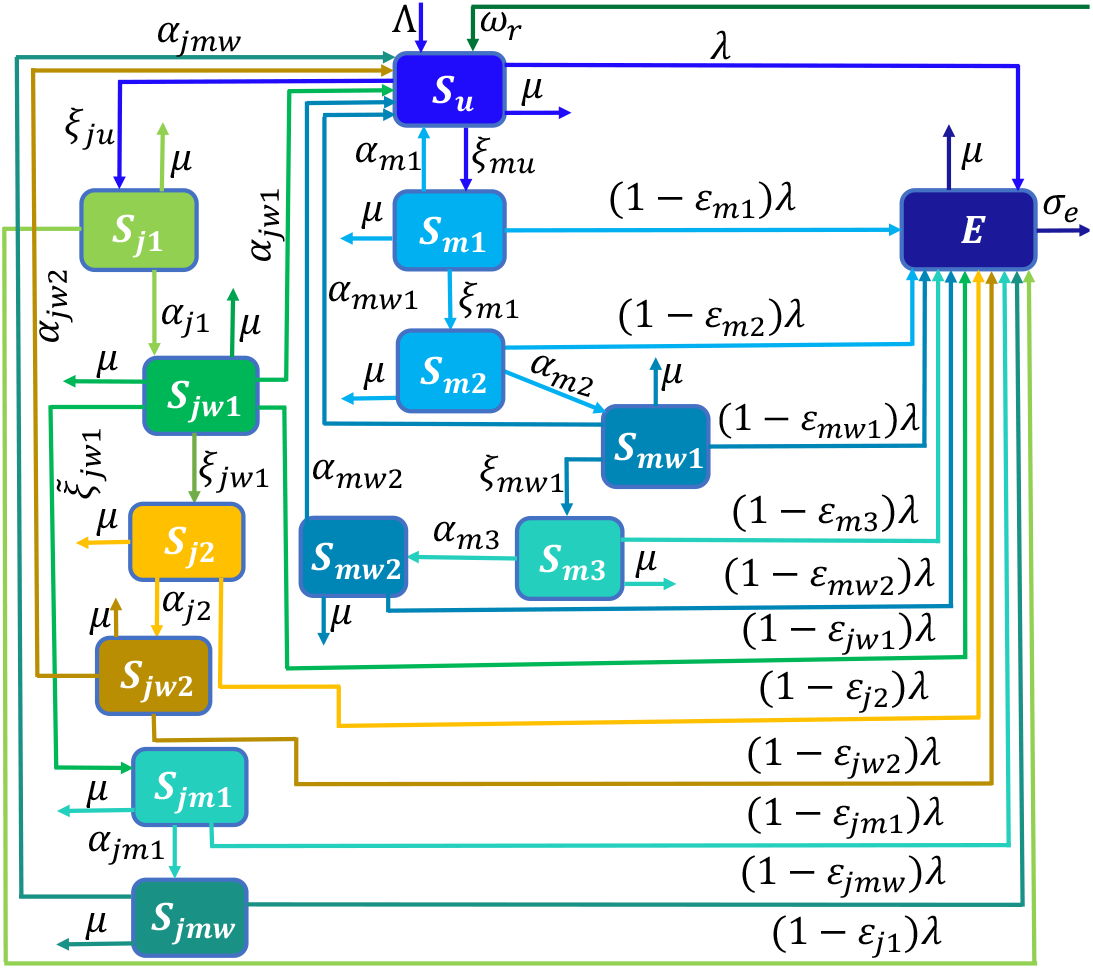
Schematics of Model 3, i.e., the vaccination model with the first booster dose. Individuals in the waned vaccine-induced immunity class (*S*_*jw*1_) join the *S*_*j* 2_ (*S*_*jm*1_) class if boosted with the J & J (mRNA) vaccine, while individuals in the *S*_*mw*1_ class join the *S*_*m*3_ class upon boosting with an mRNA vaccine. When vaccine-induced immunity wanes, individuals from the *S*_*j* 2_ (*S*_*jm*1_) class join the *S*_*j* 2_ (*S*_*jm*1_) class, while individuals from the *S*_*m*3_ class join the *S*_*mw* 2_ class. The other variables are as defined above and in Table S1 of the SI, and the rest of the schematics is as in Fig. 1. The model parameters are described in the text and Table S2 of the SI.

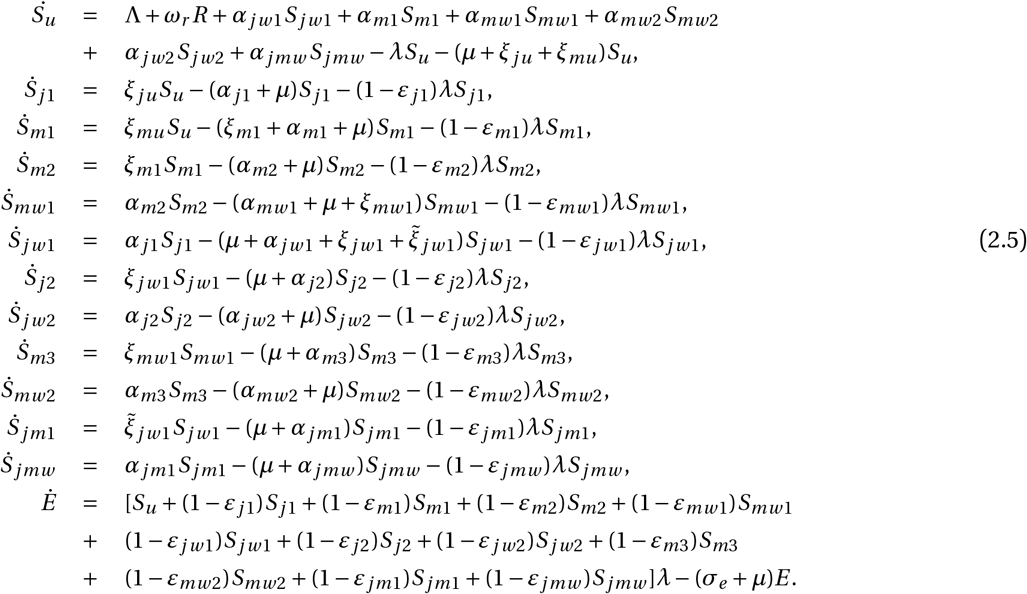

### 2.5. Model 4: The vaccination model with two booster vaccine doses

In this Section, the vaccination model with the first booster vaccine dose given by Eqs. (2.2) and (2.5) is extended to include a second booster vaccine dose. That is, individuals in the waned vaccine-induced immunity class (*S*_*jw* 2_) who were fully vaccinated and boosted with the J & J vaccine receive another dose of the J & J vaccine and progress to the class of individuals who have received a total of three J & J vaccine doses (*S*_*j* 3_) at per capita rate *ξ*_*jw* 2_, or are now boosted with an mRNA vaccine and progress to the *S*_*j* 2*m*_ class at per capita rate 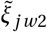. Individuals in the waned vaccine-induced immunity class (*S*_*jmw*_) who were fully vaccinated with the J & J vaccine but boosted with an mRNA vaccine receive another booster dose of an mRNA vaccine and progress to the *S*_*jm*2_ class at per capita rate *ξ*_*jmw*_. Individuals in the waned vaccine-induced immunity class (*S*_*mw* 2_) who were fully vaccinated and boosted with the same mRNA vaccine receive another booster dose of an mRNA vaccine and progress to the *S*_*m*4_ class at per capita rate *ξ*_*mw* 2_. Natural deaths in each of these classes are at per capita rate (*μ*). Individuals in the *S*_*j* 3_ (*S*_*m*4_) class progress to the *S*_*jw* 2_ (*S*_*mw* 2_) class at rate *α*_*j* 3_ (*α*_*m*4_), when their immunity wanes, while individuals in the *S*_*jm*2_ (*S*_*j* 2*m*_) class progress to the *S*_*jmw*_ class at per capita rate *α*_*jm*2_ (*α*_*j* 2*m*_), when their immunity wanes. Break-through infections in these new classes of individuals who have received a second booster vaccine dose are at rate (1−*ε*_*l*_)*λ, l* ∈ { *j* 3, *jm*2, *j* 2*m, m*4}, where 0 ≥ *ε*_*l*_ ≥ 1 are the efficacies of the second booster vaccine doses in preventing humans from being infected and *λ* is as defined in Eq. (2.1). The other variables and parameters of the model are as defined in Sections 2.2-2.4 and Tables S1 and S2 of the SI, and the total population (*N*) is given by *N* = *S*_*u*_ + *S*_*j*1_ + *S*_*m*1_ + *S*_*m*2_ + *S*_*mw*1_ + *S*_*jw*1_ + *S*_*j* 2_ + *S*_*jm*1_ + *S*_*jw* 2_ + *S*_*m*3_ + *S*_*mw* 2_ + *S*_*jmw*_ + *S*_*j* 3_ + *S*_*jm*2_ + *S*_*j* 2*m*_ + *S*_*m*4_ + *E* + *I*_*p*_ + *I*_*a*_ + *I*_*s*_ + *I*_*c*_ + *I*_*h*_ + *R*. Schematics of the unvaccinated susceptible, vaccinated, boosted, and exposed portions of the model are presented in Fig. 4, the model equations are given in (2.6), and the full vaccination model with two booster vaccine doses is described by the system of equations (2.2) and (2.6).

**Fig. 4:**
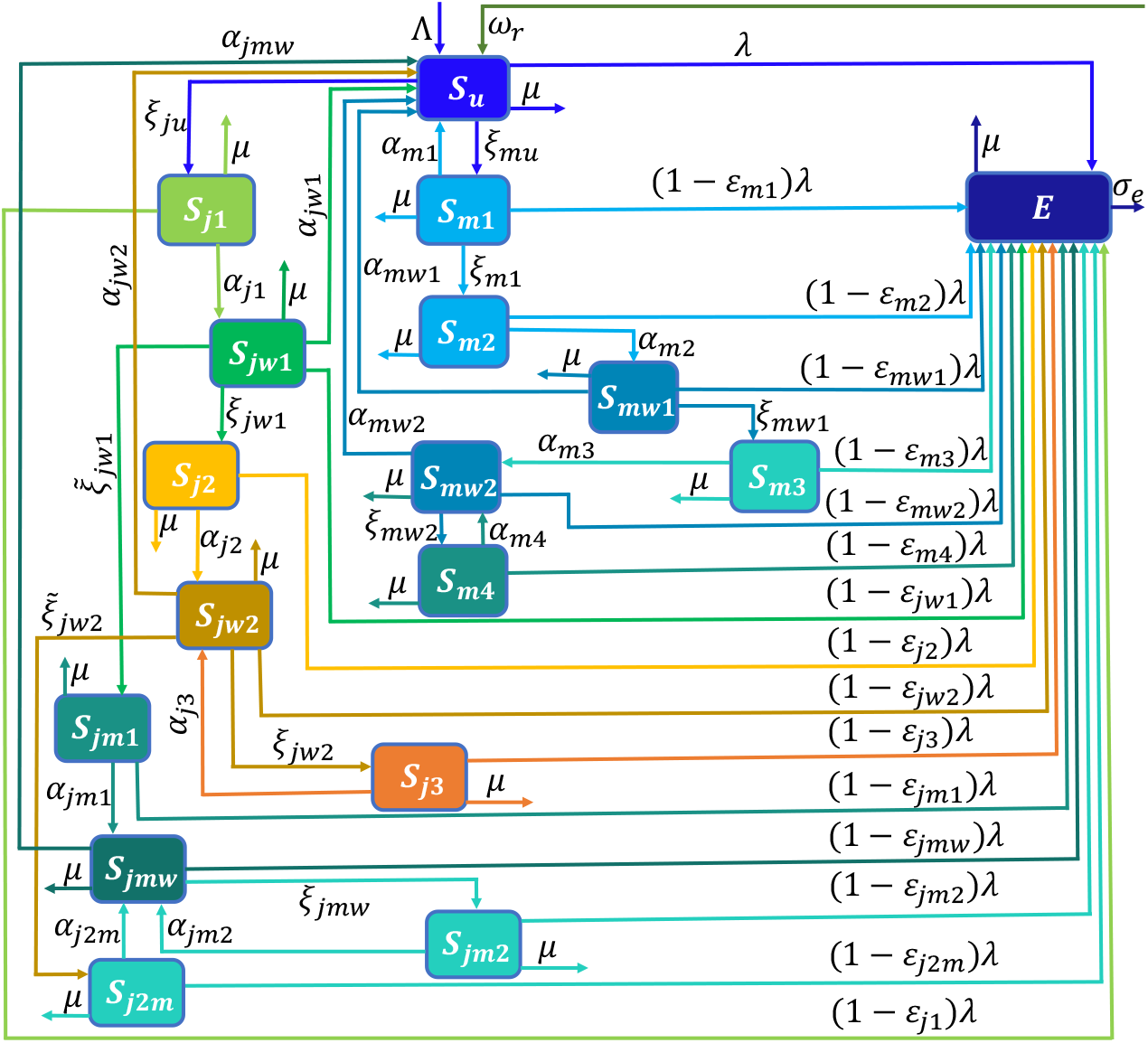
Conceptual framework of the model with two booster vaccine doses (Model 4). The first booster dose classes consist of individuals who were fully vaccinated with a J & J (mRNA) vaccine in the waned vaccine-induced immunity class *S*_*jw*1_ (*S*_*mw*1_) and then boosted with a J & J (mRNA) vaccine denoted by *S*_*j* 2_ (*S*_*j* 3_) and the class of individuals who were fully vaccinated with a J & J vaccine, but boosted with an mRNA vaccine (*S*_*jm*1_). The second booster dose classes consist of individuals who received a J & J (mRNA) first booster in the waned vaccine-induced immunity class *S*_*jw* 2_ (*S*_*jwm*_), who receive a second J & J (mRNA) booster dose denoted by *S*_*j* 3_ (*S*_*jm*2_), the class of individuals who received a J & J first booster dose, but an mRNA second booster dose denoted by (*S*_*j* 2*m*_), and the class of individuals who were fully vaccinated and boosted with an mRNA vaccine in the waned vaccine-induced class (*S*_*mw* 2_), who received a second mRNA booster dose (*S*_*m*4_). The rest of the schematics, i.e., the portion from the exposed class (*E*) through the recovered class (not shown here), is as in Fig. 1. The other classes are as defined in Sections 2.2-2.4 and described in Table S1 of the SI, while the parameters are described in text and Table S2 of the SI.

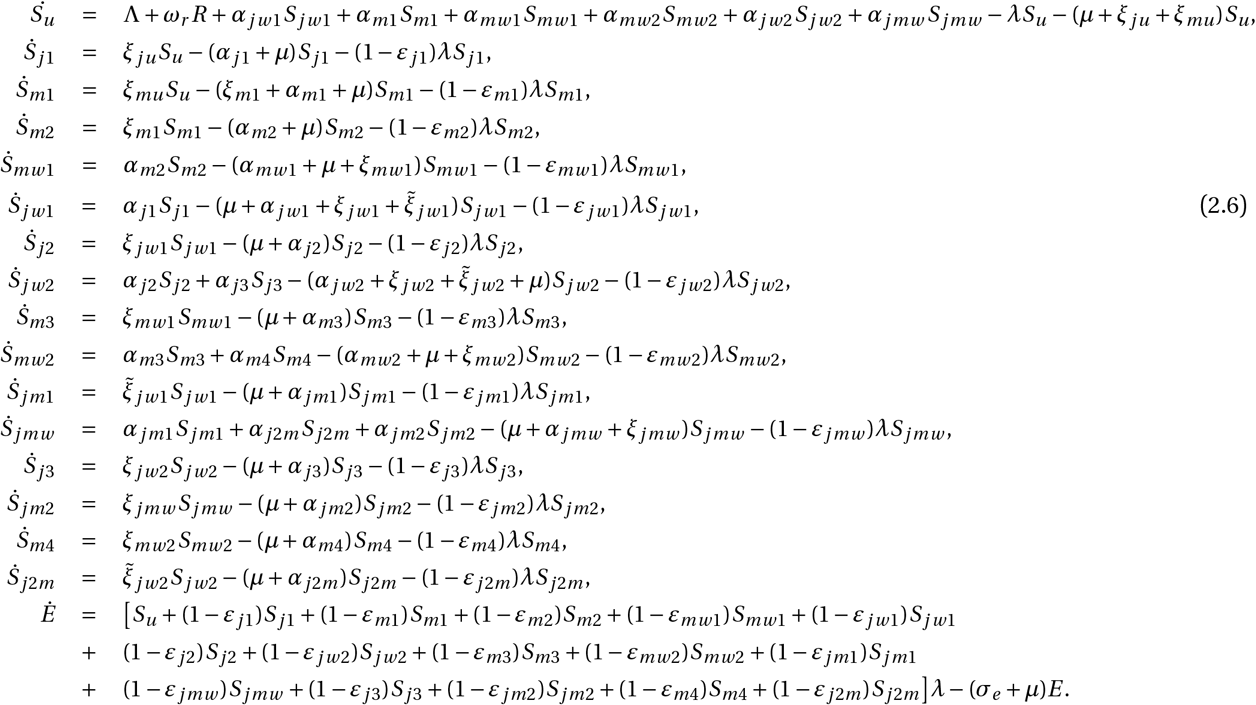

## 3. Results

### 3.1. Analytical results

In this section, we compute the disease-free equilibria and the basic and control reproduction numbers of the models developed in Section 2 and establish the stability of the disease-free equilibria. In particular, we show that the disease-free equilibrium of each model is locally asymptotically stable when the associated reproduction number is less than unity and that under specific parameter regimes, the disease-free equilibrium of each model can be globally asymptotically stable. Furthermore, we compute the endemic equilibrium of the basic model (Eqs. (2.2)-(2.3)) explicitly.

#### 3.1.1. Disease-free equilibrium

The disease-free equilibria of Models 1-4 are obtained by setting the left-hand sides of the equations of each of the models and the disease-related terms to zero. This leads to the equilibrium 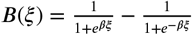, for Model 1 (Eqs. (2.2) and (2.3)) and 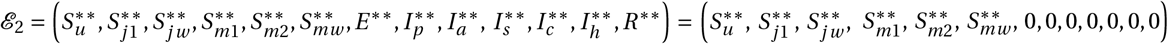, where 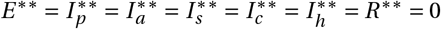 and

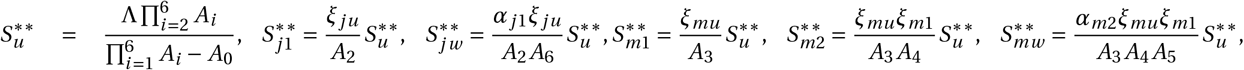

for the vaccination model (Eqs. (2.3) and (2.4)). Here, *A*_0_ = *ξ*_*mu*_ (*α*_*m*2_*α*_*mw*1_*ξ*_*m*1_ + *α*_*m*1_ *A*_4_ *A*_5_)*A*_2_ *A*_6_ + *α*_*j*1_*α*_*jw*1_*ξ*_*ju*_ *A*_3_ *A*_4_ *A*_5_, *A*_1_ = *ξ*_*ju*_ + *ξ*_*mu*_ + *μ, A*_2_ = *α*_*j*1_ + *μ, A*_3_ = *α*_*m*1_ + *ξ*_*m*1_ + *μ, A*_4_ = *α*_*m*2_ + *μ, A*_5_ = *α*_*mw*1_ + *μ, A*_6_ = *α*_*jw*1_ + *μ*, and it can be verified that the denominator of 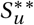 is positive (since the three negative terms in *A*_0_ are contained in the product of the *A*_*i*_ ‘s).

The disease-free equilibrium of Model 3 (Eqs. (2.3) and (2.5)) is given by 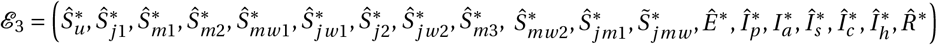, where 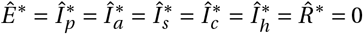, and

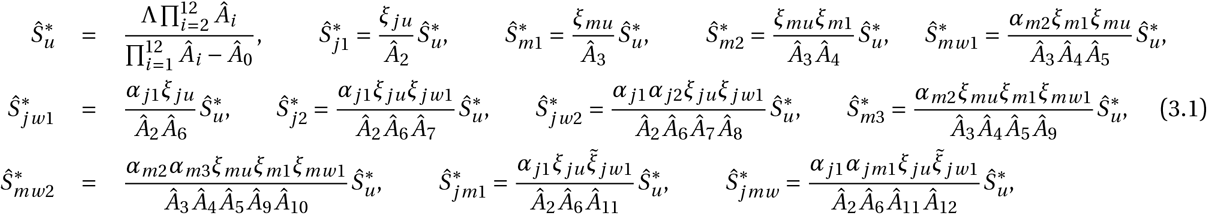

with 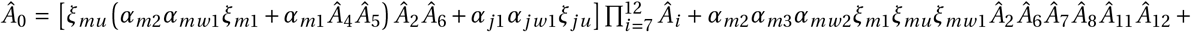 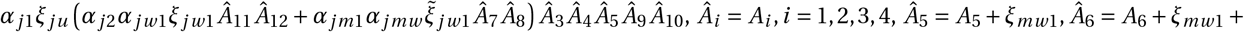 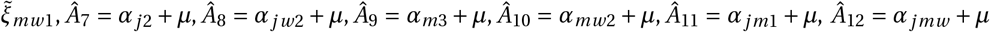. It can easily be verified that 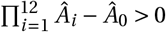 since all the terms in *Â*_0_ are contained in the product 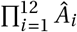.

The disease-free equilibrium of Model 4 (Eqs. (2.3) and (2.6)) is 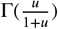, where 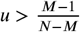, and

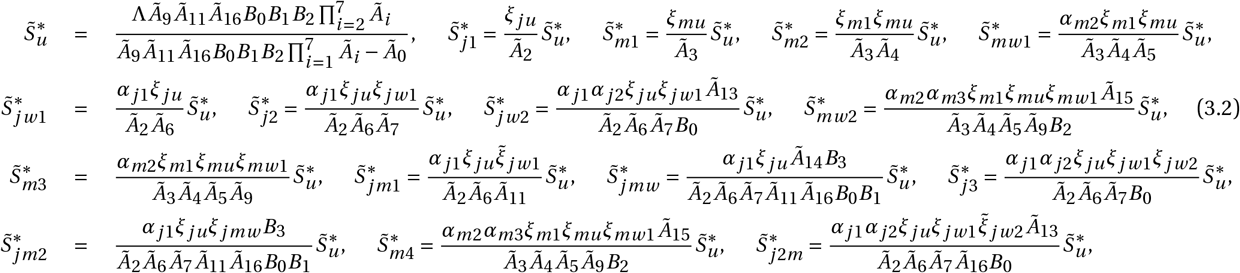

with 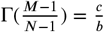 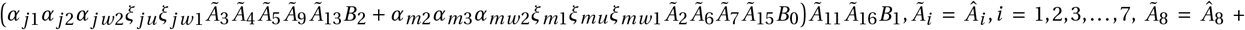 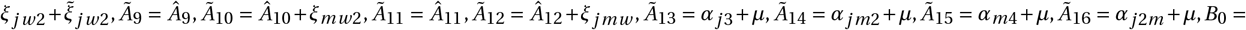 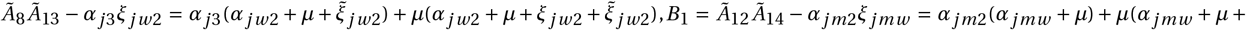 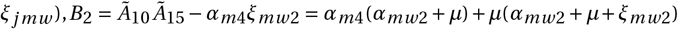, and 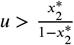. It can easily be verified that the denominator 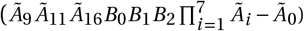 of 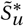 is positive since all the terms in *Ã*_0_ are contained in the product 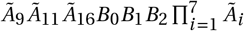.

#### 3.1.2. Reproduction number, stability of disease-free equilibria, and herd immunity threshold

The next generation operator method [86, 92] can be used to establish the asymptotic stability of the disease-free equilibria and to compute the reproduction number of Models 1-4. In particular, using the notation in [86], the matrices of new infections (*F*_*j*_, *j* = 1, 2, 3, 4) and transitions (*V*_*j*_, *j* = 1, 2, 3, 4), where *j* corresponds to the model number is given by

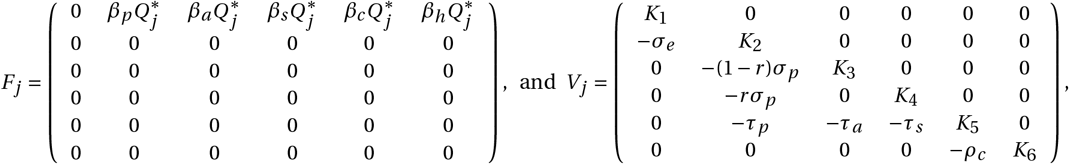

with *K*_1_ = *σ*_*e*_ + *μ, K*_2_ = *τ*_*p*_ + *σ*_*p*_ + *μ, K*_3_ = *τ*_*a*_ + *γ*_*a*_ + *μ, K*_4_ = *τ*_*s*_ + *γ*_*s*_ + *δ*_*s*_ + *μ, K*_5_ = *ρ*_*c*_ + *γ*_*c*_ + *δ*_*c*_ + *μ*, and *K*_6_ = *γ*_*h*_ + *δ*_*h*_ + *μ*. The inverse of the transfer matrix (*V*) denoted by *V*_−1_ and the next generation matrix denoted by *FV* ^−1^ are

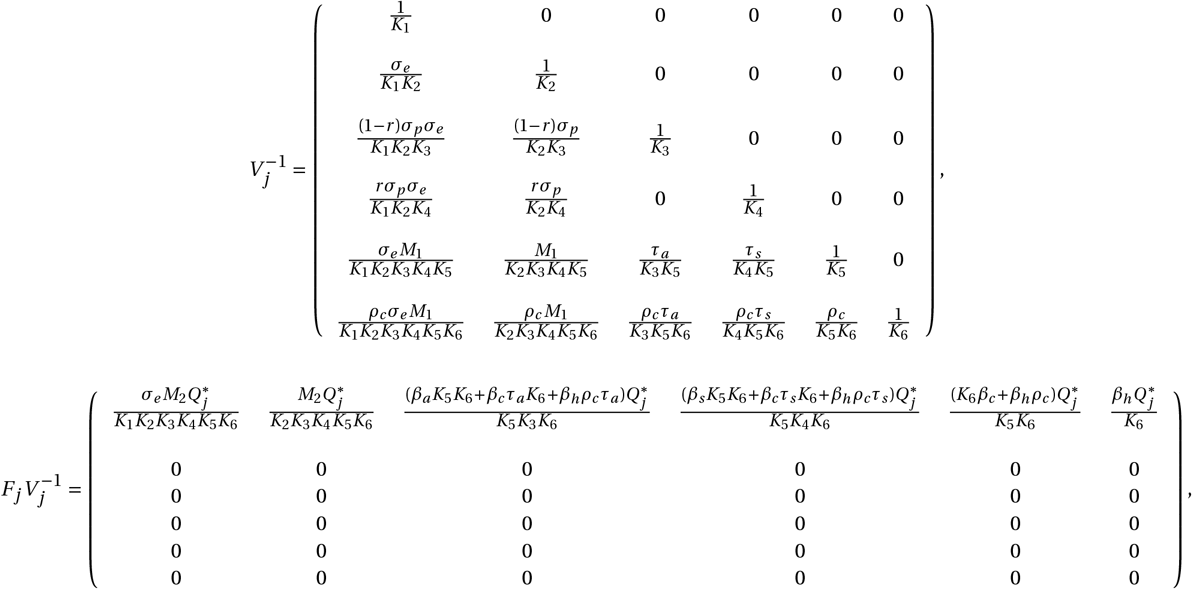

where *M*_1_ = *τ*_*p*_ *K*_3_*K*_4_ + *σ*_*p*_ [*τ*_*a*_ (1 − *r*)*K*_4_ + *rτ*_*s*_ *K*_3_], *M*_2_ = [*β*_*p*_ *K*_3_*K*_4_ + *β*_*a*_ (1 − *r*)*σ*_*p*_ *K*_4_ + *β*_*s*_*rσ*_*p*_ *K*_3_] *K*_5_*K*_6_ + (*β*_*c*_ *K*_6_ + *β*_*h*_*ρ*_*c*_)*M*_1_. In the next generation matrix 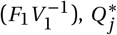 depends on the specific model under consideration. Specifically, for the basic model (i.e., Eqs. (2.2)-(2.3)), *j* = 1 and 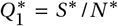; for the vaccination model with no boosting (Eqs. (2.3) and (2.4)), *j* = 2 and 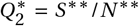, where 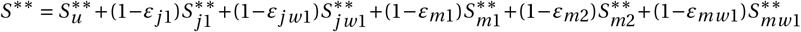 and 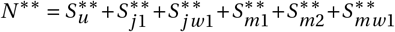; for the vaccination model with one booster dose (Eqs. (2.3) and (2.5)), *j* = 3 and 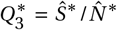, where 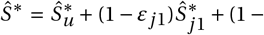 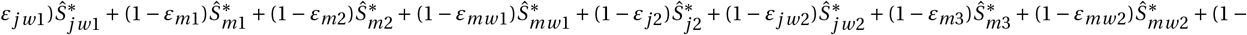 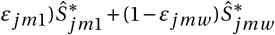 and 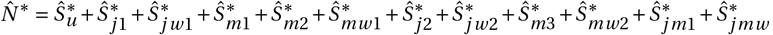; and for the vaccination model with two booster vaccine doses, *j* = 4 and 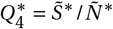, where 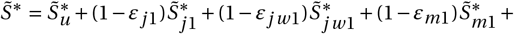 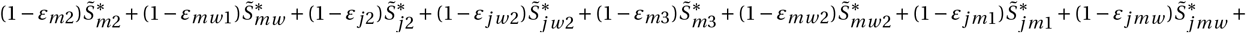 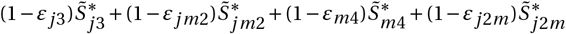 and 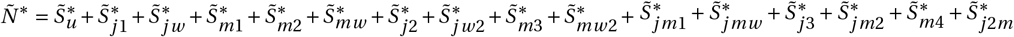.

For the four models, it is convenient to define the quantity ℛ_*c j*_, *j* = 1, 2, 3, 4, by:

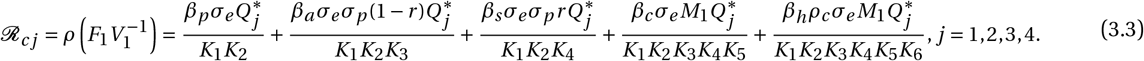

Depending on the value of 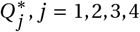, the quantity ℛ_*c j*_, is the *control or vaccination reproduction number* of Model 1 (when *j* = 1), Model 2 (when *j* = 2), Model 3 (when *j* = 3), and Model 4 (when *j* = 4). It measures the average number of new COVID-19 cases generated by a typical infectious individual introduced into a population where a certain fraction is protected throughout the period within which the individual is infectious. Mathematically, it is the spectral radius of the next generation matrix 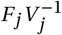. Observe that ℛ_*c j*_ is the sum of the reproduction numbers connected with disease transmission by pre-symptomatic infectious (*I*_*p*_), asymptomatic infectious (*I*_*a*_), symptomatic infectious (*I*_*s*_), confirmed (*I*_*c*_), and hospitalized (*I*_*h*_) individuals. It should be mentioned that in the absence of vaccination and any other control measures, ℛ_*c*_ reduces to the basic reproduction number (ℛ_0_), where

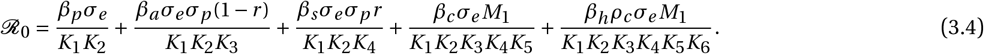

Thus, 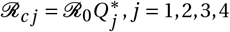. From Theorem 2 in [86], we have the following result:

##### Theorem 3.1.

*For a value of* 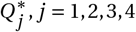 *that corresponds to any of the models in Section 2, the associated disease-free equilibrium is locally-asymptotically stable if ℛ*_*c j*_ < 1 *and unstable if ℛ*_*c j*_ > 1.

Theorem 3.1 can be interpreted, epidemiologically, to mean that a small influx of COVID-19 cases will not generate a COVID-19 out-break if the control reproduction number (ℛ_*c j*_) is less than unity. In what follows, the reproduction number of Eqs. (2.2) and (2.3), Eqs. (2.3) and (2.4), Eqs. (2.3) and (2.5), and Eqs. (2.3) and (2.6)) will be denoted by ℛ_*c*1_, ℛ_*c*2_, ℛ_*c*3_, and ℛ_*c*4_, respectively.

In addition to the reproduction number, the vaccine-induced herd immunity threshold (i.e., the minimum percentage of the population that must be immunized with an anti-COVID vaccine to stop the COVID-19 outbreak) is another important epidemiological quantity. We derive an equation from which this threshold can be determined. Let 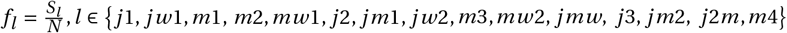, be the fraction of the susceptible (fully vaccinated) population that is required to be fully vaccinated (boosted) with an anti-COVID vaccine to attain vaccine-induced herd immunity, then the control or vaccine reproduction number 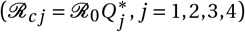 can be re-written as ℛ_*c j*_ = (1 − Σ_*l*_ *ε*_*l*_ *f*_*l*_*) ℛ*_0_. Setting ℛ_*cj*_ = 1, we have

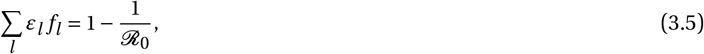

from which the herd immunity threshold (denoted by 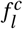) for various scenarios can be determined.

#### 3.1.3. Endemic equilibrium and global stability analysis of the basic model

Generally, equilibria of the basic model (Eqs. (2.2)-(2.3)) are obtained by setting the left hand side of the system to zero and solving the resulting system in terms of the equilibrium value (*λ*^*^) of the force of infection (*λ*). This leads to

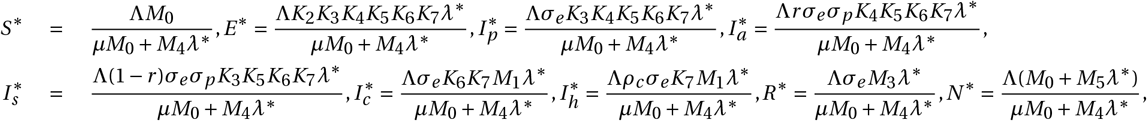

where *M*_0_ = *K*_1_*K*_2_*K*_3_*K*_4_*K*_5_*K*_6_*K*_7_, *M*_3_ = [(1−*r*)*γ*_*a*_ *K*_4_ +*rγ*_*s*_ *K*_3_]*σ*_*p*_ *K*_5_*K*_6_ +(*γ*_*c*_ *K*_6_ +*γ*_*h*_*ρ*_*c*_)*M*_1_, *M*_4_ = *M*_0_ −*ω*_*r*_ *σ*_*e*_ *M*_3_, and *M*_5_ = {(*σ*_*e*_ +*K*_2_)+[*r K*_3_ + (1 − *r*)*K*_4_]*σ*_*e*_*σ*_*p*_ }*K*_5_*K*_6_*K*_7_ + *σ*_*e*_ [(*ρ*_*c*_ + *K*_6_)*K*_7_ *M*_1_ + *M*_3_]. It can be verified that *M*_4_ = *M*_0_ − *ω*_*r*_ *σ*_*e*_ *M*_3_ > 0. Substituting 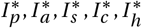 in the equilibrium expression for *λ*^*^ (obtained from Eq. (2.1)) and solving for *λ*^*^ leads to *λ*^*^ = 0, which reduces 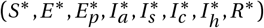 to the disease-free equilibrium (Λ/*μ*, 0, 0, 0, 0, 0, 0, 0), or *λ*^*^ = Λ*M*_0_(ℛ_*c*1_ − 1)/*M*_5_, which leads to the following result:

##### Theorem 3.2.

*The basic model (Eqs*. (2.2)*-*(2.3)*) has a unique endemic equilibrium* 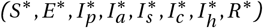, *if ℛ*_*c*1_ > 1 *and no endemic equilibrium otherwise*.

Next, we establish the global stability of the disease-free equilibrium (*E*_1_) of the model (2.2)-(2.3). Consider the positively invariant and attracting region 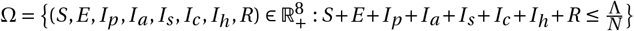, for the model (2.2)-(2.3), and the Lyapunov function: ℒ = *g*_1_*E* + *g*_2_ *I*_*p*_ + *g*_3_ *I*_*a*_ + *g*_4_ *I*_*s*_ + *g*_5_ *I*_*c*_ + *g*_6_ *I*_*h*_, where, 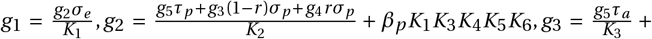 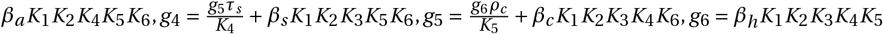. The Lyapunov derivative 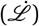 is:

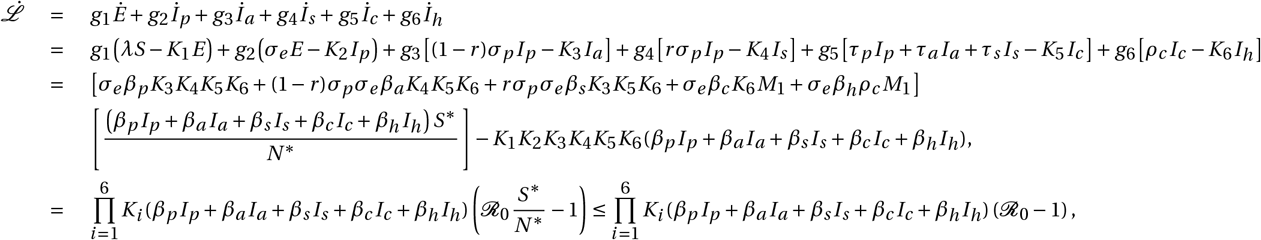

since *S*^*^ ≥ *N* ^*^. Hence, 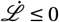 if *R*_0_ ≥ 1, and 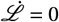 if and only if *I*_*p*_ = *I*_*a*_ = *I*_*s*_ = *I*_*c*_ = *I*_*h*_ = 0. Substituting *I*_*p*_ = *I*_*a*_ = *I*_*s*_ = *I*_*c*_ = *I*_*h*_ = 0 in the model (2.2)-(2.3) shows that 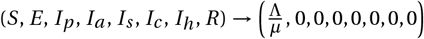, as *t* → ∞. Furthermore, it can be shown that the largest compact invariant set in 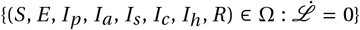 is the disease-free equilibrium of the basic model (ℰ_1_). It follows, from LaSalle’s Invariance Principle, that the disease-free equilibrium of the model (2.2)-(2.3) is globally-asymptotically stable in Ω whenever ℛ_0_ ≥ 1. This proves the following theorem:

##### Theorem 3.3.

*The disease-free equilibrium (ℰ*_1_*) of the model* (2.2)*-*(2.3) *is globally-asymptotically stable in* Ω *if ℛ*_0_ ≥ 1.

### 3.2. Data sources and parameter estimation

The four models derived in Sections 2.2-2.5 have several parameters, some of which are available in the literature or can be calculated using COVID-19 and demographic information available in the literature (see Table S3 in the SI). The remaining unknown parameters are estimated by fitting specific models to confirmed new daily case and mortality data for the U.S. from January 22, 2020, to June 13, 2022, extracted from [41, 43]. Raw (daily) case and mortality data instead of cumulative data are used for the fitting to minimize common estimation errors in the calibrated parameter values, and their confidence intervals [44]. The fitting is performed using a nonlinear least squares technique in MATLAB version R2022a. This entails identifying the best set of parameters that minimizes the sum of the square differences given by 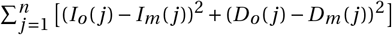, where *I*_*o*_ (*D*_*o*_) are the observed confirmed new COVID-19 cases (deaths) from the data and *I*_*m*_ = *τ*_*p*_ *I*_*p*_ + *τ*_*a*_ *I*_*a*_ + *τ*_*s*_ *I*_*s*_ (*D*_*m*_ = *δ*_*s*_ *I*_*s*_ + *δ*_*c*_ *I*_*c*_ + *δ*_*h*_ *I*_*h*_) are the daily cases (deaths) from the model [25, 54, 55, 96]. Minimization of the sum of the squared differences is accomplished with the “*lsqcurvefit*” function in MATLAB, which takes input matrices of confirmed daily cases and deaths from the observed data and model, as well as bounds and an initial guess for the parameter values and outputs the best set of estimated parameters and residuals (that can be used to compute confidence intervals) among other outputs. The 95% confidence intervals of the estimated parameter values are obtained through a bootstrap method with 10, 000 bootstrap samples (see [25, 45] for details). Since the entire data set from January 22, 2020, to June 13, 2022, includes different waves and events that occurred at different times, the model fitting is based on different waves, events, and/or policies implemented during the COVID-19 era. Specifically, the entire data set is split into 8 periods consisting of: i) the first COVID-19 wave (January 22 to June 1, 2020); ii) the second COVID-19 wave (June 2 to September 14, 2020); iii) the first part of the third COVID-19 wave, i.e., the period from September 15, 2020, to the onset of vaccination (December 19, 2020); iv) the second part of the third COVID-19 wave, i.e., the period from the onset of vaccination (December 19, 2020) to July 4, 2021; v) the first portion of the fourth COVID-19 wave, i.e., the period from July 5 to September 24, 2021 (just before the onset of boosting); vi) the period from the start of boosting to the onset of the Omicron variant of concern (September 25 to December 2, 2021); vii) the period of the main Omicron wave to the start of second booster shots (December 3, 2021, to March 29, 2022); and viii) the period from the start of the second booster shots to June 13, 2022. The basic model (2.2)-(2.3) is fitted to the data segments from January 22 to December 18, 2020, while the vaccination model with no booster (Eqs. (2.3) and (2.4)) is fitted to the data segment from December 19, 2020, to September 22, 2021. The vaccination model with the first booster dose (Eqs. (2.3) and (2.5)) is fitted to the data segments from September 25 to December 2, 2021, and December 3, 2021, to March 28, 2022, while the vaccination model with two booster doses (Eqs. (2.3) and (2.6)) is fitted to the data segment from March 29 to June 13, 2022. The rest of the data (i.e., the segment from June 14 to December 11, 2022) is used to validate or illustrate the model’s performance. Additionally, both cumulative case and mortality data are used to validate the models. Results of the model fit and validation are depicted in Fig. 5 (a)-(b) and Fig. 5 (c)-(d), respectively. The validation results show an excellent match between the model and the remaining daily data (light green curves in Fig. 5 (a) and (c)), and a perfect match between the cumulative cases and deaths from the model and the cumulative case and mortality data for the period from January 22, 2020, to December 11, 2022 (Fig. 5 (b) and (d)). The estimated parameter values and their associated 95% confidence intervals are reported in Tables S4-S8 of the SI, while the initial conditions used are reported in Section 3 of the SI.

**Fig. 5:**
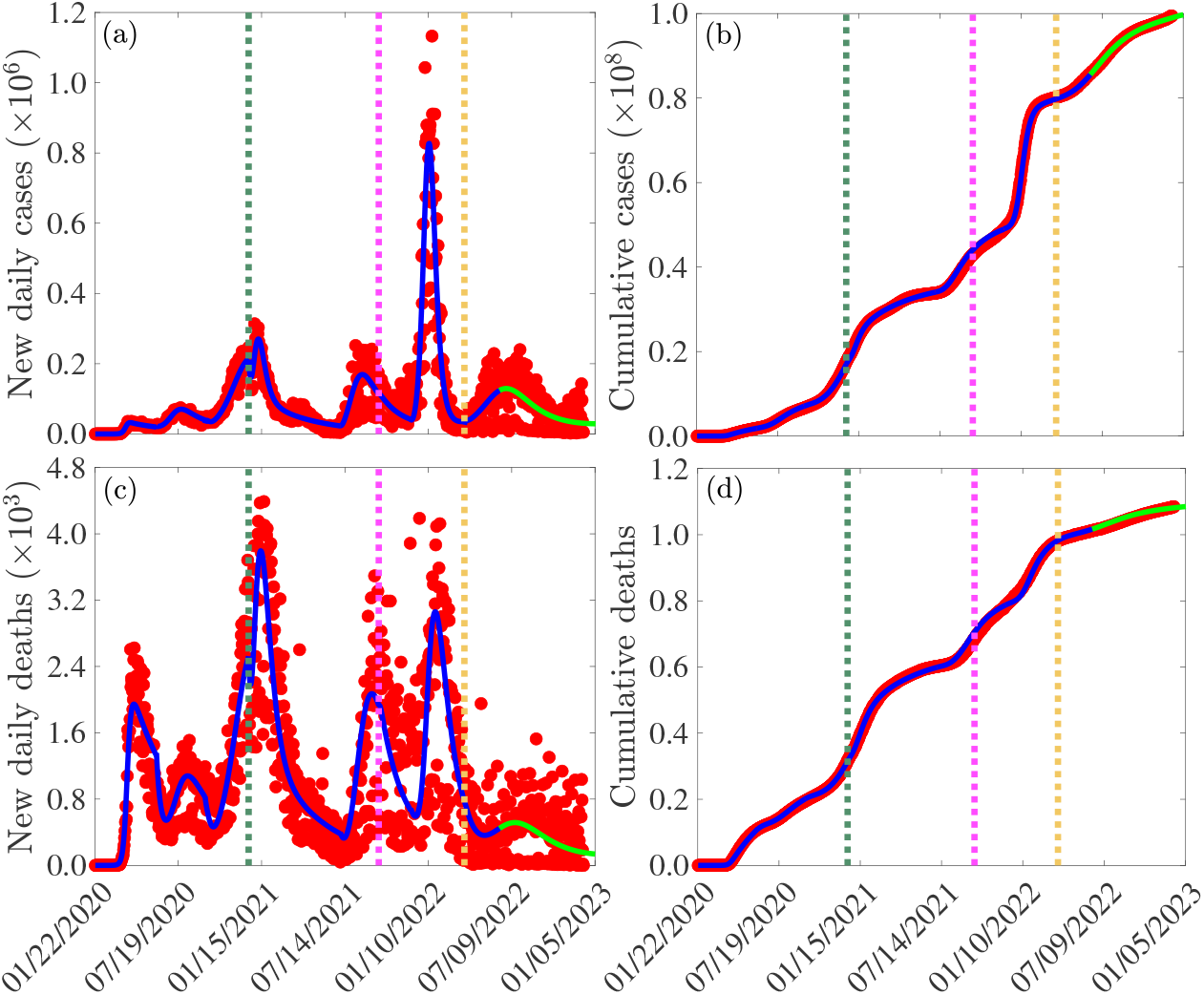
Fitting of the models given by Eqs. (2.2) and (2.3) (January 22 to December 18, 2020), Eqs. (2.3) and (2.4) (December 19, 2020, to September 24, 2021), Eqs. (2.3) and (2.5) (September 25, 2021, to March 28, 2022), and Eqs. (2.3) and (2.6) (March 29, 2022, to June 13, 2022) to (a) new daily COVID-19 cases and (c) new daily COVID-19 mortality data for the United States. The performance of the models is illustrated by simulating the models using the estimated parameters and plotting the cumulative (b) case and (d) mortality outputs of the models and the corresponding cumulative data. Dotted dark green, magenta, and gold vertical lines depict the start of vaccination, first booster, and second booster, respectively, while light green line segments (from June 14, 2022, to December 11, 2022) show validation of the fitted model. The fixed and estimated parameter values are presented in Tables S3-S8 of the SI, while the initial conditions are presented in Section 3 of the SI.

Fitting of the model to data reveals that the transmission rate of COVID-19 in the U.S. was highest during the original Omicron wave, which is consistent with observed data [37]. Also, the fitting shows that apart from the Delta and Omicron waves, the transmission rate was highest during the first wave (i.e., from January 22 - June 1, 2020). This high transmission rate can be explained by the lack of, poor adherence to, or ineffectiveness of non-pharmaceutical Interventions (NPIs) implemented during this period. The control reproduction number during this period was ℛ_*c*1_ ≈ 3.66 with confidence interval (1.89, 4.83). Relaxation of lockdown and other control measures, as well as events such as the July 4, 2021 celebration, caused a surge in the number of cases which resulted in the second wave of the pandemic [62]. This wave was driven primarily by pre-symptomatic and asymptomatic infectious individuals, as was the case with the first wave [50, 62–64]. In particular, our estimated parameters indicate that pre-symptomatic and asymptomatic infectious individuals were the main drivers COVID-19 transmission in the U.S. Inconsistent use of NPIs during the post-lockdown period and mass testing resulted in an average positivity rate of 1.4% [43, 51] contributed to the surge that resulted in the big pandemic wave from September 15 to December 18, 2020. The control reproduction number for this period was ℛ_*c*1_ = 4.94 with confidence interval (2.68, 5.38) before the start of vaccination and ℛ_*c*2_ = 0.94 with confidence interval (0.69, 1.18) during the initial vaccination period (from December 19, 2020, to July 4, 2021). Hence, emergency-use authorization of COVID-19 vaccines in the U.S. that were highly effective in preventing severe disease and symptomatic infections [16, 20, 28] was timely and useful in curtailing the burden of this wave. Despite the availability of these highly effective vaccines, another surge in cases, which resulted in an 89% increase in the reproduction number from the previous period, was witnessed from July 5 to September 24, 2021. This surge was attributed to factors such as vaccine hesitancy [52], waning of natural and vaccine-induced immunity [53, 56, 58], non-compliance with NPIs [62], and the emergence of the Delta variant of concern (VOC), which was more transmissible than the wild-type virus [59]. The next surge from early December 2021 was attributed to the Omicron VOC, which was more transmissible than any other variant, although it resulted mostly in milder cases, fewer hospitalizations, and fewer deaths [60, 61, 101–103]. Specifically, our model fitting estimates the peak number of cases (deaths) during the Omicron wave to be 827, 025 (2, 830) compared to 169, 054 (2, 069) cases (deaths) during the previous period. That is, the number of cases (deaths) at the peak of the Omicron wave was about 5 (1.4) times the number of cases (deaths) during the previous wave. Also, the cumulative number of cases (deaths) during the Omicron wave was about 30, 879, 500 (181, 522), which is ≈ 2 (0.95) times the cumulative number of cases (deaths) during the previous wave. The reproduction number for this period is ℛ_*c*3_ = 3.59 with confidence interval (2.14, 5.76), which is ≈ 3.6 times the reproduction number of the previous period. This is consistent with reports in [23]. Finally, the reproduction number for the wave that occurred after the Omicron wave is ℛ_*c*4_ = 1.76 with confidence interval (0.85, 2.25). Despite the relaxation of NPIs, the reproduction number of this wave is about half that of the Omicron wave, and our model estimates the number of cases (deaths) at the peak of this wave to be 0.16 (0.18) times the number of cases (deaths) at the peak of the Omicron wave. This significant reduction can be attributed to the increase in boosting vaccine-induced immunity among fully vaccinated individuals.

#### 3.2.1. Threshold vaccination level

The baseline parameter values in Tables S3-S8 of the SI and the herd immunity equation (3.5) are used to compute the herd immunity thresholds under different vaccine efficacy scenarios. In particular, if the population is fully vaccinated only with an mRNA (J & J) vaccine with an average efficacy of 94.55%(67%), the herd immunity threshold is ≈ 71.6% (100%), while if the population is fully vaccinated with a combination of mRNA and J & J vaccines, the herd immunity threshold is ≈ 79%. This threshold can go as high as 96% if the waning of vaccine-induced immunity is considered. Since a combination of these vaccines are used in the U.S., and ≈ 68% of the U.S. populace were fully vaccinated by early December 2022 [19], an additional 11% of the U.S. populace is required to be vaccinated to attain vaccine-induced herd immunity if it is assumed that vaccine-induced immunity does not wane. However, since it has been established that vaccine-induced immunity wanes over time [3, 48], fully vaccinating (boosting) a sizeable proportion of the unvaccinated (fully vaccinated but not boosted) population might be necessary for achieving herd immunity.

Figure 6 depicts elaborate profiles of the control reproduction number (ℛ_*c*_) of Model 2 (ℛ_*c*_ = ℛ_*c*2_), Model 3 (ℛ_*c*_ = ℛ_*c*3_), and Model 4 (ℛ_*c*_ = ℛ_*c*4_) as functions o{f vaccine efficacy (*ε*_*v*_) and}the proportion (*f*_*v*_) of the population that is vaccinated/boosted. It should be noted that for Model 2, {*ε*_*v*_ ∈{*ε*_*j*1_, *ε*_*jw*1_, *ε*_*m*1_, *ε*_*m*2_, *ε*_*mw* 1}_; for Model 3, *ε*_*v*_ ∈ {*ε*_*j*1_, *ε*_*jw*1_, *ε*_*m*1_, *ε*_*m*2_, *ε*_*mw*1_, *ε*_*m*3_, *ε*_*mw* 2_, *ε*_*j* 2_, *ε*_*jw* 2_, *ε*_*jm*1_, *ε*_*jmw*_}, and for Model 4, *ε*_*v*_ ∈ {*ε*_*j*1_, *ε*_*jw*1_, *ε*_*m*1_, *ε*_*m*2_, *ε*_*mw*1_, *ε*_*m*3_, *ε*_*mw* 2_, *ε*_*j* 2_, *ε*_*jw* 2_, *ε*_*jm*1_, *ε*_*jmw*_, *ε*_*m*4_, *ε*_*j* 3_, *ε*_*jm*2_, *ε*_*j* 2*m*}_. Vaccine-induced herd immunity thresholds associated with various vaccine efficacies can be derived from these contour plots. For example, in the absence of boosting, ≈ 71.2% (71.9%) of the population is required to be fully vaccinated to attain vaccine-induced herd immunity if only the Pfizer (Moderna) vaccine with an efficacy of 95% (94.1%) is used (Fig. 6 (a)). These thresholds are higher if some people, who receive the first dose of these mRNA vaccines fail to return for the second dose and when vaccine-induced immunity wanes over time. In particular, if vaccine-induced immunity wanes to a level at which the average protective efficacy of the vaccine falls to 75% (67%), then ≈ 90% (≈ 100%) of the population must be vaccinated to achieve vaccine-induced herd immunity. Thus, if vaccine-induced immunity wanes to a level at which the average protective efficacy of the vaccine falls below 67%, then reducing the control reproduction number below unity without boosting is unattainable even if the entire population is vaccinated. For the case in which vaccine-induced immunity wanes and fully vaccinated individuals receive only the first booster dose, ≈ 85% of the population is required to be fully vaccinated and boosted to attain vaccine-induced herd immunity if only mRNA vaccines are used, while ≈ 94% of the population is required to be fully vaccinated and boosted to attain vaccine-induced herd immunity if a combination of the J & J and mRNA vaccines are used (Fig. 6 (b)). Furthermore, if vaccine-induced immunity wanes over time and fully vaccinated individuals are boosted twice, ≈ 87% of the population is required to be fully vaccinated and boosted to attain vaccine-induced herd immunity provided only mRNA vaccines are used, while ≈ 96% of the population is required to be fully vaccinated and boosted to attain vaccine-induced herd immunity if both the J & J and mRNA vaccines are used (Fig. 6 (c)). Hence, this study shows that, even with the highly effective three vaccines against COVID-19 that are authorized for use in the U.S., a substantial fraction of the population is required to be fully vaccinated and boosted to attain vaccine-induced herd immunity in the U.S., if waning of vaccine-induced immunity is fast.

**Fig. 6:**
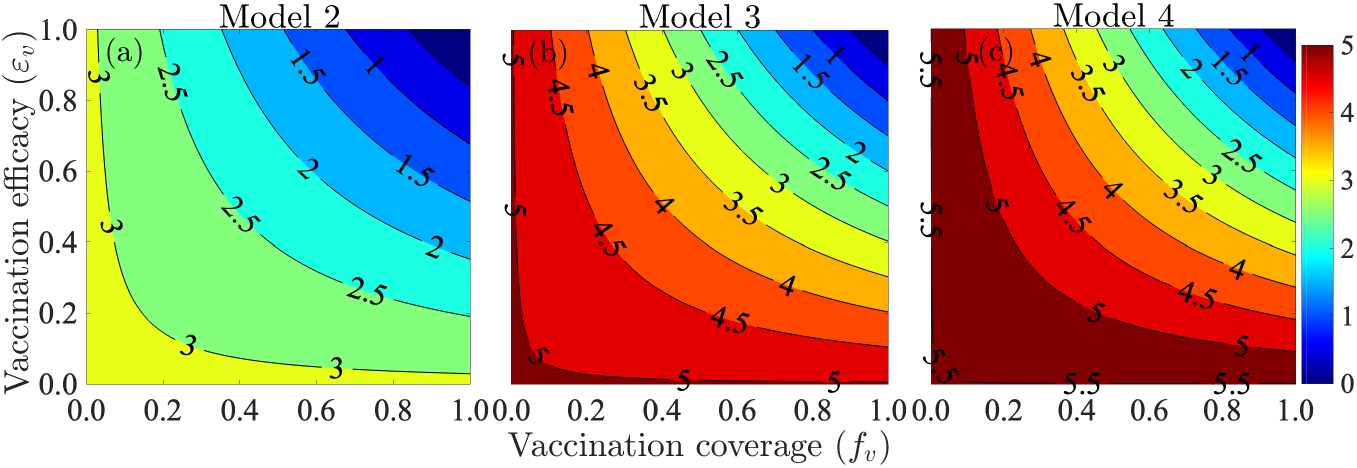
Contour plot depicting the profile of the control reproduction number (ℛ_*c*_) of (a) Model 2 (ℛ_*c*2_), (b) Model 3 (ℛ_*c*3_), and (c) Model 4 (ℛ_*c*4_) as a function of vaccine efficacy (*ε*_*v*_) and the vaccinated proportion of the population (*f*_*v*_). For Model 2, *ε*_*v*_ ∈ {*ε*_*j*1_, *ε*_*jw*1_, *ε*_*m*1_, *ε*_*m*2_, *ε*_*mw*1_ ; for Model 3, *ε*_*v*_ ∈ {*ε*_*j*1_, *ε*_*jw*1_, *ε*_*m*1_, *ε*_*m*2_, *ε*_*mw*1_, *ε*_*m*3_, *ε*}_*mw* 2_, *ε*_*j* 2_, *ε*_*jw* 2_, *ε*_*jm*1_, *ε*_*jmw* }_, and for Model 4, *ε*_*v*_ ∈ {*ε*_*j*1_, *ε*_*jw*1_, *ε*_*m*1_, *ε*_*m*2_, *ε*_*mw*1_, *ε*_*m*3_, *ε*_*mw* 2_, *ε*_*j* 2_, *ε*_*jw* 2_, *ε*_*jm*1_, *ε*_*jmw*_, *ε*_*m*4_, *ε*_*j* 3_, *ε*_*jm*2_, *ε*_*j* 2*m* }_. The values of the parameters used for producing this plot are given in Tables S3, S6 (a), S7 (b), and S8 of the SI.

### 3.3. Numerical simulation results

In this section, the four models derived in Sections 2.2-2.5 are simulated using the fixed parameter values in Table S3 and the estimated parameter values in Tables S4-S8 to assess the impact of 1) the type of vaccine (J & J only, mRNA, or a combination of J & J and mRNA vaccines) used for vaccination and boosting; 2) timing (early versus late implementation) of vaccination and boosting; 3) mass vaccination and boosting; 4) relaxing and reinforcing vaccination and transmission reducing measures such as masking-up; and 5) the impact of waning natural and vaccine-induced immunity on the burden of the COVID-19 pandemic in the U.S. In all the graphs presented in this section, the baseline case (using the parameters in Tables S3-S8 of the SI) is denoted by a blue curve.

#### 3.3.1. Assessing the impact of the type of vaccine used for vaccination and boosting

To investigate the impact of the single-dose J & J or any of the two-dose mRNA vaccines on COVID-19 in the U.S., Models 2, 3, and 4 are simulated using the parameters in Tables S3-S8 of the SI. The results of the simulation (presented in Fig. 7) show that the baseline number of daily cases (deaths) peaked by January 8, 2021 (January 12, 2021) with a peak size of ≈ 230, 785 (≈ 3, 548) cases (deaths) during the wave in which vaccination started in the U.S. (blue curves in Fig. 7 (a) and (d)), August 27, 2021 (September 16, 2021) with a peak size of 158, 913 cases and 2, 071 deaths during the Delta wave (blue curves in Fig. 7 (b) and (e)), January 12, 2022 (January 27, 2022) with 818, 223 cases and 2, 860 deaths at the peak of the Omicron wave, and June 29, 2022 (July 18, 2022) with a peak size of 123, 270 cases and 495 deaths during the wave in which the second booster uptake started (blue curves in Fig. 7 (c) and (f)). The study shows that for the worst-case scenario in which no vaccination program was implemented, a 150% and 158% increase in the baseline peak number of daily cases and deaths, respectively, would have been recorded during the wave in which vaccination started and a 208% (144%) increase in the baseline peak number of daily cases (deaths), would have been recorded during the Delta wave (comparing the peaks of the blue and magenta curves in Fig. 7 (a) and (d)). Also, the study shows that for the worst-case scenario in which no boosting program was implemented, a 50% (38%) increase in the baseline peak number of daily cases (deaths), would have been recorded during the Delta wave (comparing the peaks of the blue and magenta curves in Fig. 7 (b) and (e)), while a 9% (8%) increase in the baseline peak number of daily cases (deaths) would have been recorded during wave in which the second booster uptake started, if no second booster program was adopted (comparing the peaks of the blue and magenta curves in Fig. 7 (c) and (f)).

**Fig. 7:**
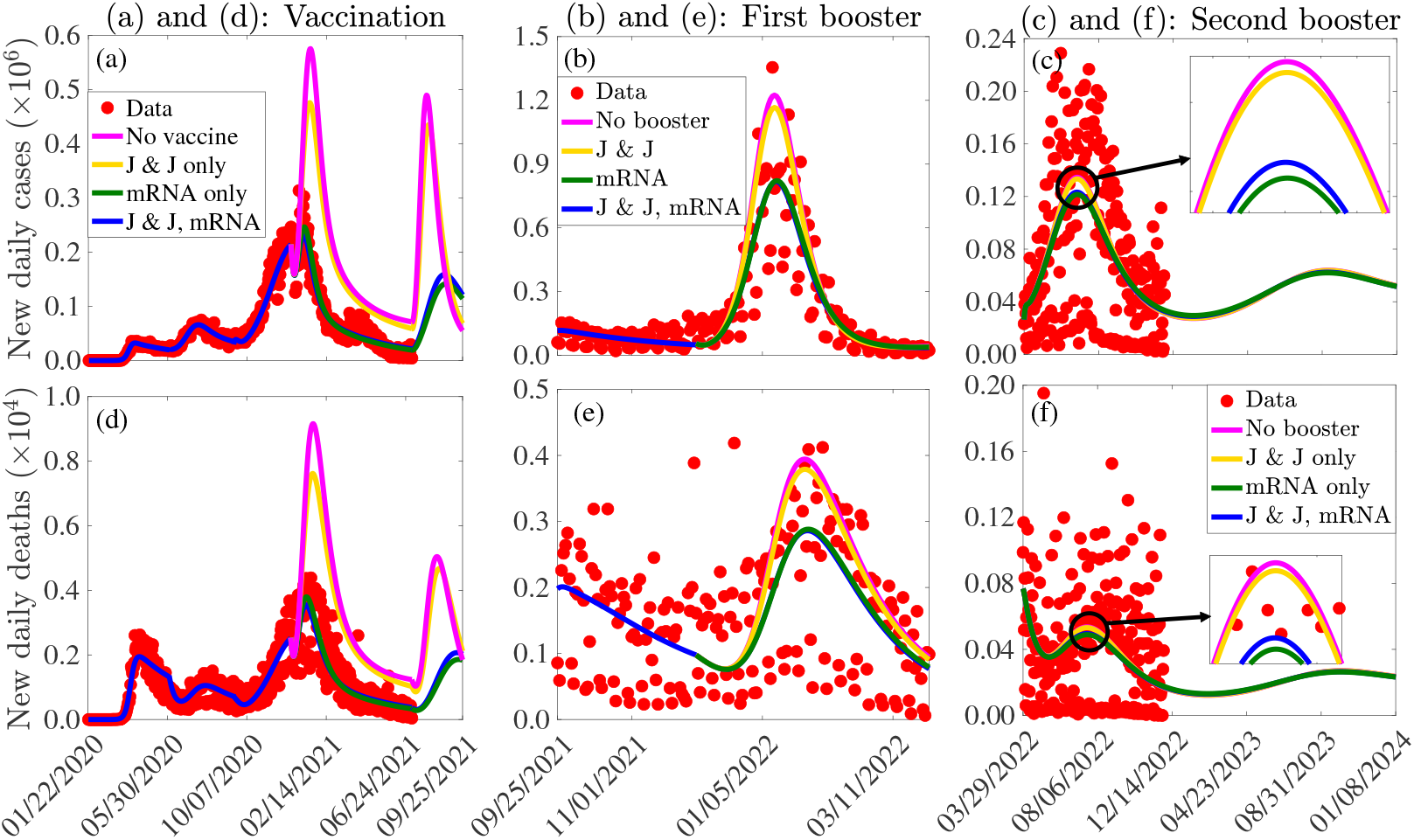
Simulations of Models 1-4 in Sections 2.2-2.5 depicting the impact on new daily cases ((a)-(c)) and deaths ((d)-(f)) of using the J & J vaccine (magenta curves), the Pfizer-BioNTech or Moderna (mRNA) vaccine (dark green curves), or both the J & J and mRNA vaccines (blue curves) for vaccinating unvaccinated individuals ((a) and (d)), first booster uptake for fully vaccinated individuals ((b) and (e)), and for second booster shots ((c) and (f)). The zoomed-in windows highlight the difference in peak sizes for various scenarios. The other parameters used for the simulations are given in Tables S3-S8.

If only the single-dose J & J vaccine with an efficacy of ≈ 67% was prioritized for full vaccination and boosting, then a 106% (114%) increase in the baseline peak number of daily cases (deaths) would have been registered during the wave in which vaccination started and a 174% (126%) increase in the baseline peak number of daily cases (deaths) would have been recorded during the Delta wave (comparing the peaks of the blue and gold curves in Fig. 7 (a) and (d)). Furthermore, if only the J & J vaccine was used for vaccination and first booster uptake, a 43% (33%) increase in the baseline peak number of daily cases (deaths) would have been recorded during the Omicron wave (comparing the peaks of the blue and gold curves in Fig. 7 (b) and (e)), while an 8% (7.5%) increase in the baseline peak number of daily cases (deaths) would have been registered during the wave in which the second booster uptake started (comparing the peaks of the blue and gold curves in Fig. 7 (c) and (f)). On the other hand, if only mRNA vaccines (with an average efficacy of ≈ 94.55%) were used for vaccination and boosting, ≈ 11% (10%) of the baseline peak number of daily cases (deaths) would have been averted during the wave in which vaccination started (comparing the peaks of the blue and dark green curves in Fig. 7 (a) and (d)), while ≈ 1.4% (1.2%) of the baseline peak number of daily cases (deaths) would have been averted during the wave in which second booster uptake started (comparing the peaks of the blue and dark green curves in Fig. 7 (c) and (e)).

Similar changes are observed with the cumulative number of cases and deaths (Fig. S1 in the SI). Specifically, if no vaccination program was adopted, a 67% (74%) increase from the baseline number of cumulative cases (deaths) would have been recorded by September 25, 2021 (comparing the blue and magenta curves in Fig. S1 (a) and (d)). If a vaccination program was adopted, but no booster uptake was adopted, the baseline number of cumulative cases (deaths) would have increased by 12% (5%) by March 29, 2022 (comparing the blue and magenta curves in Fig. S1 (b) and (e)), while if a full vaccination and first booster program were adopted, but with no second booster program, the baseline number of cumulative cases (deaths) will increase by 0.7% (0.3%) by the end of 2023 (comparing the blue and magenta curves in Fig. S1 (c) and (f)). If only the J & J vaccine had been administered from the start of vaccination in the U.S., a 56% (59%) increase in the number of cumulative cases (deaths) would have been recorded by September 25, 2021 (comparing the blue and gold curves in Fig. S1 (a) and (d)). Lower increases in the cumulative number of cases and deaths compared to the worst-case scenario would have been recorded if only the J & J vaccine was used for boosting (comparing the blue and gold curves in Fig. S1 (b)-(c) and (e)-(f)). A vaccine program that prioritized only mRNA vaccines from the start of vaccination in the U.S. would have resulted in a reduction of 1.6% (1.4%) in the reported number of cumulative cases (deaths) by September 25, 2021 (comparing the blue and green curves in Fig. S1 (a) and (d)). This represents an ≈ 69% (75%) reduction in comparison to the worst-case scenario with no vaccination (comparing the magenta and green curves in Fig. S1 (a) and (d)). Similar reductions would have been obtained if only mRNA vaccines were used for boosting.

#### 3.3.2. Assessing the impact of vaccination and booster timing

The models developed in Sections 2.2-2.5 are simulated to assess the impact of timing of vaccination and booster doses on the burden of COVID-19 (quantified in terms of the number of confirmed cases and deaths) in the U.S. Specifically, we evaluate the impact of starting vaccination and boosting two or three weeks earlier, and then two weeks later than the actual date on the dynamics of COVID-19 in the U.S. The results obtained and depicted in Fig. 8 show that if vaccination in the U.S. started two weeks later (i.e., from January 1, 2021, instead of the assumed December 19, 2020), the number of new daily cases (deaths) during the wave in which vaccination started would have peaked one week earlier than the baseline scenario, with 425, 560 cases (6, 463 deaths) at the peak (gold curves in Fig. 8 (a) and (d)). This represents a drastic 95% (93%) increase from the baseline peak number of daily cases (deaths). For this hypothetical case (in which vaccination started two weeks later), a 15% (17%) increase in the cumulative number of cases (deaths) would have been recorded by January 25, 2021. The reduction in the time for the pandemic to attain a peak and the time to elimination would have been even more significant if vaccination had started earlier. In particular, if vaccination in the U.S. started two weeks earlier (i.e., on December 5, 2020), the number of cases (deaths) during the third wave of the pandemic would have peaked about 12 days earlier compared to the baseline scenario, with a 27% (24%) reduction from the baseline number of daily cases (deaths) at the peak (comparing the blue and light green curves in Fig. 8 (a) and (d)). For this scenario, ≈ 20% (17%) of the cumulative number of cases (deaths) would have been averted by January 25, 2021 (comparing the blue and light green curves in Fig. S2 (a) and (d)). Furthermore, if vaccination started three weeks earlier (i.e., from November 28, 2020), the number of cases (deaths) during the third wave of the pandemic would have peaked 18 (20) days earlier compared to the baseline case and a more significant decrease in the baseline peak number of daily cases (49%) and deaths (44%) would have been recorded (comparing the blue and dark green curves in Fig. 8 (a) and (d)). For this case, ≈ 33% (29%) of the cumulative cases (deaths) would have been averted by January 25, 2021.

**Fig. 8:**
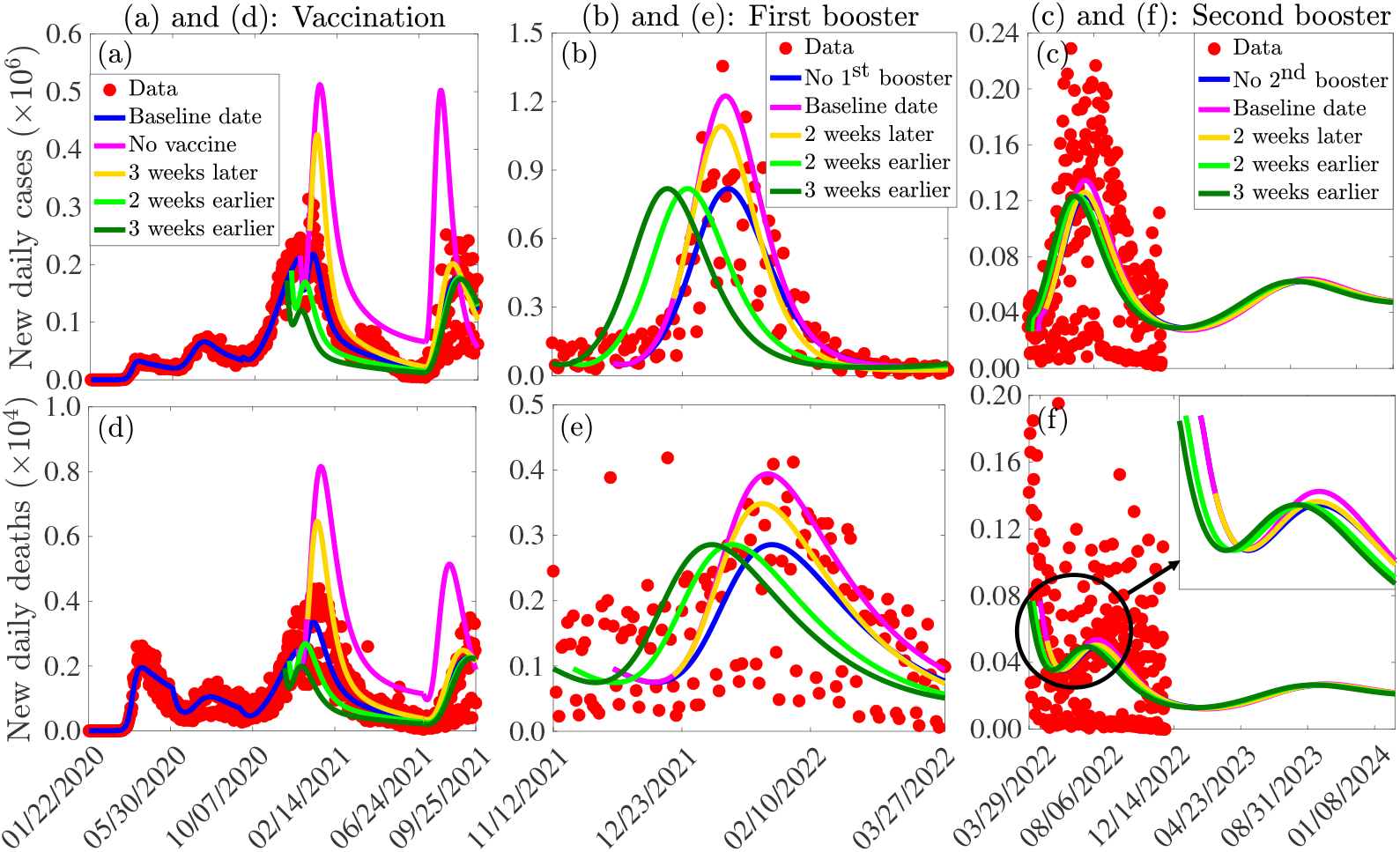
Simulations of Models 1-4 in Sections 2.2-2.5 depicting the impact of administration timing of vaccination ((a) and (d)), first booster update ((b) and (e)), and second booster uptake ((c) and (f)) on the daily number of COVID-19 cases ((a)-(c)) and the daily number of COVID-19 deaths ((d)-(f)) in the U.S. The parameters used for the simulations are presented in Tables S3-S8 in the SI.

Since the number of cases and deaths was low and declining when administration of the first booster doses started, we assess the impact of the first booster dose on the Omicron wave. If the first booster uptake started two weeks later during the Omicron wave, the number of cases (deaths) during the Omicron wave would have peaked 2 (3) days earlier compared to the baseline case, with a 34% (22%) increase in the baseline number of daily cases (deaths) at the peak (comparing the blue and gold curves in Fig. 8 (b) and (e)). Also, a 5% (2.5%) increase in the baseline number of cumulative cases (deaths) would have been recorded by March 29, 2022 (comparing the blue and gold curves in Fig. S2 (b) and (e)). Early administration of the first booster uptake would have resulted in reductions in the time for the pandemic to peak and slight decreases in the cumulative number of cases and deaths. In particular, if administration of the first booster shots started two weeks earlier, a 1.19% (1.34%) decrease from the baseline number of cumulative cases (deaths) would have been recorded by March 29, 2022 (comparing the blue and light green curves in Fig. S2 (b) and (e)), while if administration of the first booster dose started three weeks earlier, a 1.22% (1.80%) decrease from the baseline number of cumulative cases (deaths) would have been recorded by March 29, 2022 (comparing the blue and dark green curves in Fig. S2 (b) and (e)).

Administration of the second booster dose has been useful in curtailing the burden of the COVID-19 pandemic, albeit to a lower extent compared to primary vaccination and the first booster uptake. If the second booster uptake started two weeks later (i.e., on April 12 instead of March 29, 2022), a 2.2% (1.8%) increase from the baseline peak number of cases (deaths) would have been recorded (comparing the peaks of the blue and gold curves in Fig. 8 (c) and (f)). Also, a 0.07% (0.04%) increase in the baseline number of cumulative cases (deaths) will ill be recorded by the end of January 2023 (comparing the blue and gold curves in Fig. S2 (c) and (f)). On the other hand, if administration of the second booster dose started three weeks earlier (i.e., on March 8, 2022, instead of March 29, 2022), the wave in which the second booster uptake was administered would have peaked earlier (comparing the peaks of the blue and dark green curves in Fig. 8 (c) and (f)). Each of these second booster timing scenarios predicts a subsequent minor wave in which the daily cases (deaths) will peak around September 9, 2023 (October 4, 2023), with a peak size of 62, 427 (265) cases (deaths).

#### 3.3.3. Assessing the impact of vaccine and booster uptake

The models formulated in Sections 2.2-2.5 are simulated using the baseline parameter values presented in Table S3-S8 of the SI to assess the impact of vaccination, the first booster uptake by fully vaccinated individuals, and the second booster uptake by individuals who received the first booster shots earlier on the trajectory of the COVID-19 pandemic in the U.S. Here, it is assumed that vaccination in the U.S. started on December 19, 2020 (i.e., approximately a week after the Pfizer-BioNTech vaccine was authorized for emergency-use and a day after the Moderna vaccine was issued emergency-use authorization by the U.S. FDA), boosting of fully vaccinated individuals started on September 25, 2021, and that administration of second booster shots started on March 29, 2022. Also, we set 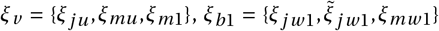, and 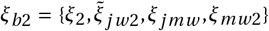. Five different vaccination and boosting scenarios are considered: i) the baseline case in which all parameters of the models are maintained at their baseline values in Tables S3-S8, ii) the worst-case scenarios in which 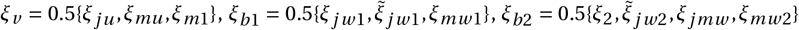, iii) the low vaccination and boosting case 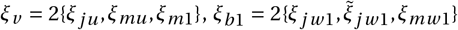, iv) the moderately high vaccination and boosting case 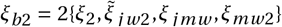, and iii) the high vaccination and boosting case 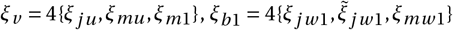, and 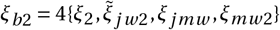.

The results of the simulation (depicted in Fig. 9) show that if all parameters of the models are held at their baseline values given in Tables S3-S8, it will be impossible to reduce the number of new daily cases or deaths below one (blue curves in Fig. 9 (a) and (d)). Hence, the disease will become endemic, i.e., establish itself in the community. Under the worst-case scenario in which no vaccination or boosting program was implemented (i.e., *ξ*_*v*_ = *ξ*_*b*1_ = *ξ*_*b*2_ = 0), it will be impossible to reduce the number of new daily cases or deaths below one. In particular, under this worst-case scenario, a 150% (158%) increase in the peak baseline number of daily cases (deaths) would have been registered in the wave in which vaccination started, and a 208% (144%) increase in the peak baseline number of daily cases (deaths) would have been registered during the Delta wave (magenta curves in Fig. 9 (a) and (d)). If a vaccination program was implemented at a rate that was 50% less than the actual rate and not complemented with booster shots (i.e., *ξ*_*v*_ ≠ 0, *ξ*_*b*1_ = *ξ*_*b*2_ = 0), a 45% (48%) increase in the peak baseline number of daily cases (deaths) would have been registered during the wave in which vaccination started and a 58% (47%) increase from the peak baseline number of daily cases (deaths) would have been registered during the Delta wave (gold curves in Fig. 9 (b) and (e)). Increasing the vaccination rate above the baseline value would have resulted in a reduction in the peak size of various waves, and a significant number of cases and deaths would have been averted. In particular, if the baseline vaccination rate was doubled, a 28% (30%) reduction in the peak baseline number of daily cases (deaths) would have been registered during the wave in which vaccination started, and a 48% (47%) reduction in the peak baseline number of daily cases (deaths) would have been registered during the Delta wave (comparing the blue and dark green curves in Fig. 9 (a) and (d)).

**Fig. 9:**
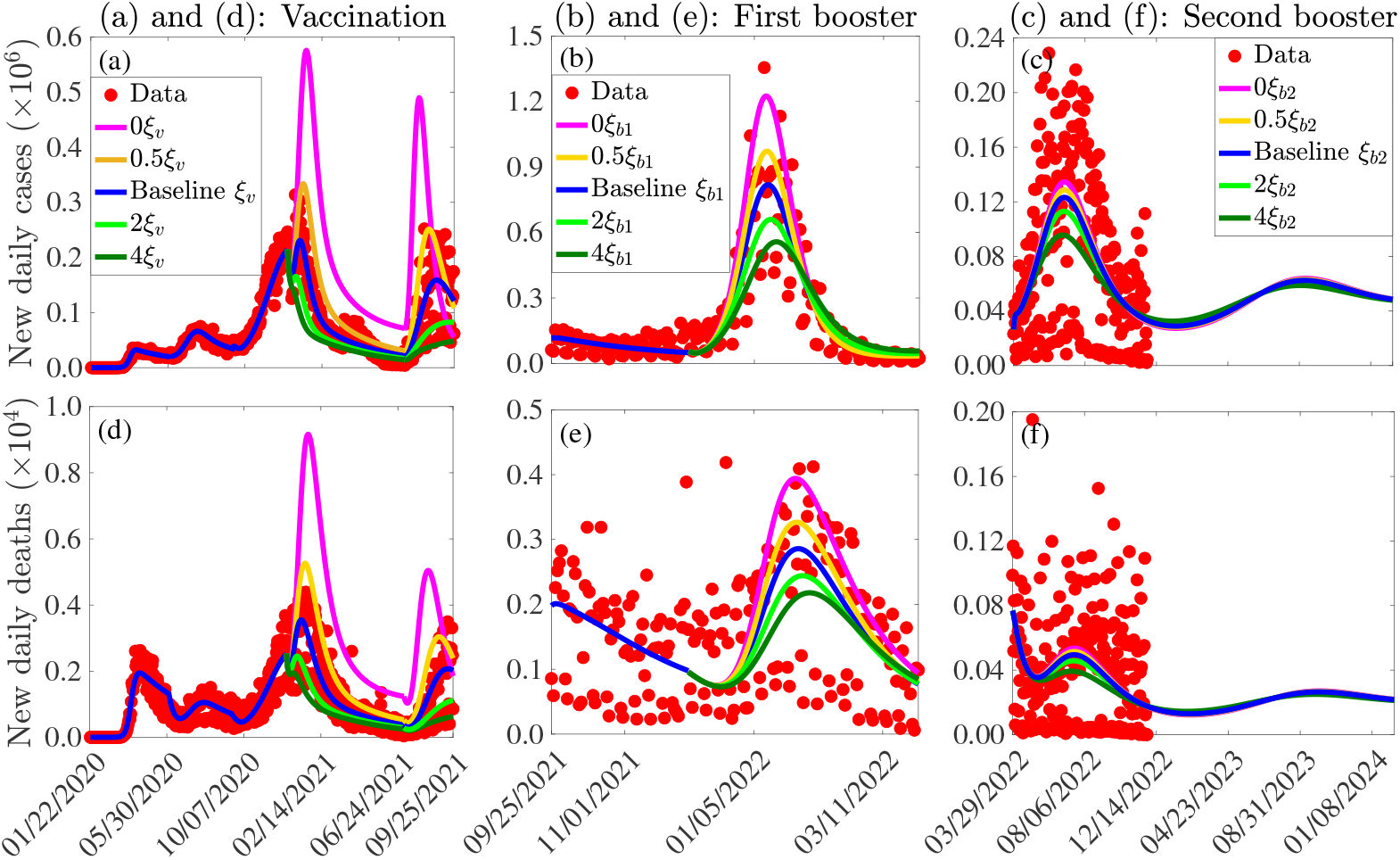
Simulations of Models 1-4 to assess the impact of vaccination ((a) and (d)), first booster uptake ((b) and (e)), and second booster uptake ((c) and (f)), on the confirmed daily cases ((a) and (c)) and deaths ((e) and (f)). The vaccination rate is *ξ*_*v*_ = {*ξ*_*ju*_, *ξ*_*mu*_, *ξ*_*m*1_}, the first booster uptake rate is 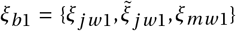, and the second booster uptake rate is (*ξ*_*b*2_) is given by 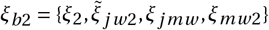. The other parameter values used for the simulations are given in Tables S3-S8.

If the vaccination program was not complemented with boosting (i.e., if *ξ*_*v*_ ≠ 0, *ξ*_*b*1_ = 0, *ξ*_*b*2_ = 0), a 50% (38%) increase from the peak baseline number of daily cases (deaths) would have been registered during the Omicron wave (comparing the peaks of the magenta and blue curves in Fig. 9 (b) and (e)). However, if a vaccination program that was complemented with a first booster shot uptake but no second booster uptake was implemented (i.e., *ξ*_*v*_ ≠ 0, *ξ*_*b*1_ ≠ 0, *ξ*_*b*2_ = 0), the increase in the peak baseline number of daily cases (deaths) would have reduced to only 19% (14%), if the vaccination and boosting rates were half of their baseline values (comparing the blue and gold curves in Fig. 9 (b) and (e)). If the first booster uptake rate was quadrupled, a 32% (24%) reduction in the peak baseline number of daily cases (deaths) would have been recorded (comparing the peaks of the blue and dark green curves in Fig. 9 (b) and (e)).

Furthermore, for a vaccination program that includes two booster uptakes after full vaccination (i.e., *ξ*_*v*_ ≠ 0, *ξ*_*b*1_ ≠ 0, *ξ*_*b*2_ ≠ 0), a 5% (4%) increase in the baseline peak number of daily cases (deaths) would have been recorded during the wave in which second boosting started if the vaccination and booster uptake rates were half of their baseline values (comparing the peaks of the blue and gold curves in Fig. 9 (c) and (f)). Accelerated boosting would have reduced the peak size of the cases and deaths. In particular, under the high boosting scenario, it would have been possible to reduce the size of the peak of the confirmed cases (deaths) by ≈ 23% (21%) during the wave in which second boosting started (comparing the peaks of the blue and dark green curves in Fig. 9 (c) and (f)). Similar changes are obtained for the cumulative cases and deaths (Fig. S3). In particular, for a vaccination program with only a first booster uptake but no second booster uptake, a 0.60% (0.25%) increase in the cumulative number of cases (deaths) will be recorded by June 30, 2023 (comparing the blue and magenta curves in Fig. S3 (c) and (f)), whereas under an accelerated vaccination program that includes two booster uptakes in which the vaccination and boosting rates are four times their baseline values, approximately 2% (1%) of the baseline cumulative cases (deaths) will be averted by June 30, 2023 (comparing the blue and dark green in Fig. S2 (c) and (f)).

#### 3.3.4. Assessing the impact of relaxing or reinforcing control measures implemented in the U.S

The model given by Eqs. (2.3) and (2.6) in section 2.5 is simulated using the fixed parameter values in Table S3 and the estimated baseline parameter values in Table S8 of SI to assess the impact of relaxing or reinforcing vaccination and transmission rate reduction control measures (such as masking-up and social distancing), as well as the impact of more transmissible new COVID-19 variants on the number of cases and COVID-related deaths in the U.S. For the case in which transmission rate reduction measures are relaxed (reinforced), the effective transmission rates (i.e., the *β*′_*j*_ *s, j* ∈ {*p, a, s, c, h*}) are multiplied by 1 + *c*_*m*_ (1 − *c*_*m*_), where 0 ≥ *c*_*m*_ ≥ 1 is the percentage increase or decrease in transmission corresponding to the level of relaxation or reinforcement. The results of these simulations (depicted in Figs. 10-11) show that in the worst-case scenario in which vaccination and boosting were completely terminated on December 1, 2022, the next (seventh) wave of the pandemic in the U.S. will attain its peak number of cases (deaths) by March 22, 2023 (April 10, 2023), with a peak size of 206, 063 confirmed new daily cases and 766 deaths (magenta curves in 10 (a) and (d)). Under this scenario, our model suggests the possibility of an eighth wave in which the number of cases (deaths) will peak about 6.6 months after the seventh wave, with the peak size of the confirmed cases (deaths) about 48% (41%) of the peak size of the worst-case scenario (comparing the second and third peaks of the magenta curves in 10 (a) and (d)), and ≈ 102% (97%) greater than the baseline peak number of cases (deaths) (comparing the second peaks of the magenta curves with the third peaks of the blue curves in 10 (a) and (d)).

**Fig. 10:**
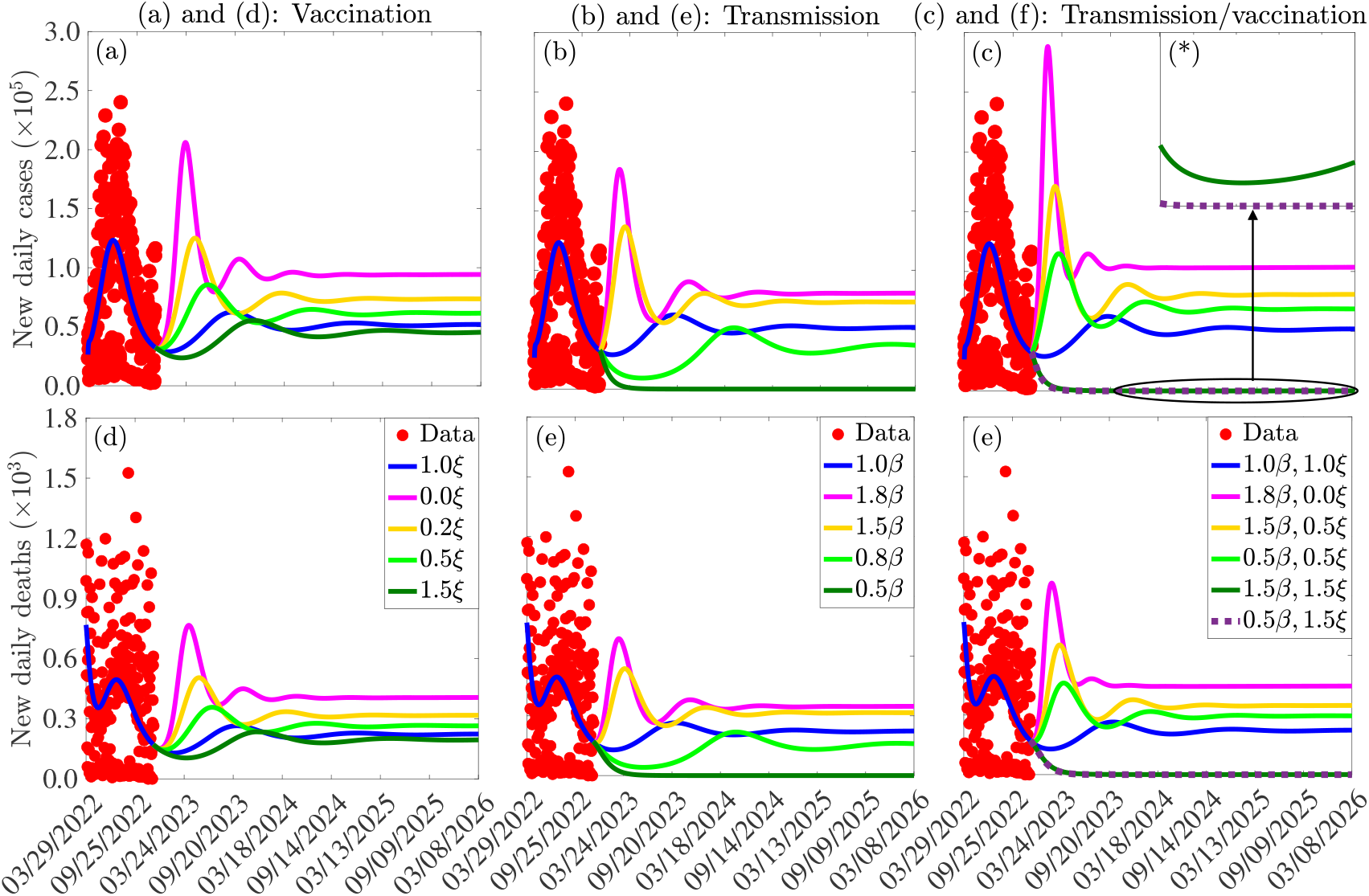
Simulations of Model 4 depicting the impact of relaxation/reinforcement in vaccination ((a) and (d)), transmission reducing control measures ((b) and (e)), and both vaccination and transmission reducing measures ((c) and (f)) on the daily confirmed COVID-19 cases ((a)-(c)) and deaths ((d)-(f)) in the U.S. The vector *ξ* entries are the baseline vaccination rates associated with mRNA and the J & J vaccines, while *β* = {*β*_*p*_, *β*_*a*_, *β*_*c*_, *β*_*c*_, *β*_*h*_ }. The other parameters used for the simulations are given in Tables S3 and S8 in the SI.

If all parameters of the model are maintained at their baseline values, the confirmed new daily cases (deaths) during the next (seventh) wave of the pandemic will peak on September 9, 2023 (October 4, 2023), with the peak sizes of the confirmed cases and deaths 62, 427 and 265, respectively, (second peaks of blue curves in 10 (a) and (d)). This represents a 223% and 189% reduction from the projected worst-case scenario number of confirmed cases and deaths, respectively, (comparing the second peaks of the blue curves with the first peaks of the magenta curves in Fig. 10 (d)). This baseline scenario represents a 15% reduction from the projected worst-case scenario number of cumulative cases and a 6% reduction from the projected worst-case scenario number of cumulative deaths by June 30, 2023 (comparing the blue and magenta curves in Fig. 11 (a) and (d)). If the number of people vaccinated and boosted per day was reduced by half on December 1, 2022, then a 99% (91%) increase from the baseline peak number of confirmed cases (deaths) will be recorded when the number of cases (deaths) during the seventh wave peak around April 18, 2023 (May 13, 2023), with the possibility of a rebound in the number of cases and deaths later (comparing the second peaks of the blue curves with the first peaks of the gold curves in Fig. 10 (a) and (d)). Compared to the projected worst-case scenario, a 38% (34%) reduction in the number of cases (deaths) at the peak of the seventh wave will be recorded (comparing the second peaks of the magenta and gold curves in Fig. 10 (a) and (d)). For this case in which the number of vaccinated and boosted individuals is halved, a 9% (3%) increase in the cumulative number of cases (deaths) will be recorded by June 30, 2023 (comparing the blue and gold curves in Fig. 11 (a) and (d)). Further reductions in the vaccination and boosting rates will lead to even more significant increases in the number of confirmed daily and cumulative cases and deaths. However, increasing the number of vaccinated and boosted people per day will reduce the daily and the cumulative number of cases and deaths. In particular, if the number of people vaccinated and boosted per day was increased by 50% on December 1, 2022, then a 11.4% (11.3%) decrease from the baseline peak number of confirmed cases (deaths) will be recorded when the seventh wave peaks, with no possibility of a significant rebound (comparing the second peaks of the blue curves and the first peaks of the dark green curves in Fig. 10 (a) and (d)). Also, a 2% (1%) decrease from the baseline number of cumulative cases (deaths) will be averted by June 30, 2023 (comparing the blue and dark green curves in Fig. 11 (a) and (d)).

**Fig. 11:**
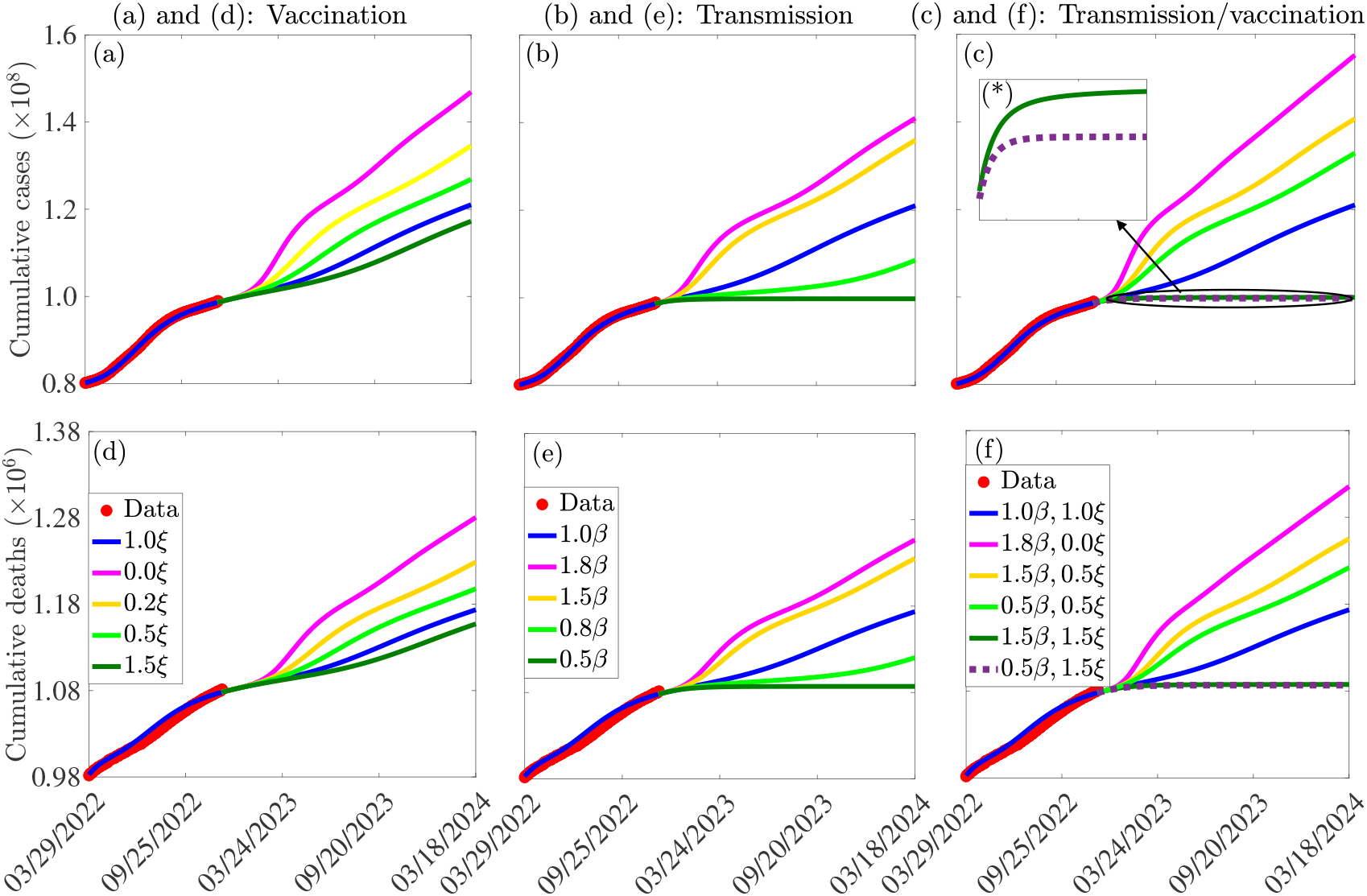
Simulations of Model 4 depicting the impact of relaxation/reinforcement in vaccination ((a) and (d)), transmission rate reducing measures ((b) and (e)), and both vaccination and transmission reducing measures ((c) and (f)) on the daily confirmed COVID-19 cases ((a)-(c)) and deaths ((d)-(f)) in the U.S. The entries of the vector *ξ* are the baseline vaccination rates associated with mRNA and the J & J vaccines, while *β* = {*β*_*p*_, *β*_*a*_, *β*_*c*_, *β*_*c*_, *β*_*h*_ }. The other parameters used for the simulations are given in Tables S3 and S8 in the SI.

On the other hand, if the vaccination and boosting rates and all other parameters are held at their baseline values and COVID-19 control measures such as mask use and social distancing that result in a reduction in disease transmission are relaxed (reinforced), a significant increase (decrease) in the number of cases and deaths can occur depending on the level of relaxation (reinforcement). Specifically, if transmission rate reduction measures were relaxed by 50% on December 1, 2022, a 119% (102%) increase in the baseline number of confirmed daily cases (deaths) will be registered when the seventh wave peaks, while an eighth wave with a smaller peak size will be registered later (comparing the second peaks of the blue curves with the first peaks of the gold curves in Fig. 10 (b) and (e)). Under this 50% reduction scenario, an 11% (4%) increase in the baseline number of cumulative cases (deaths) will be registered by June 30, 2023 (comparing the blue and gold curves in Fig. 11 (b) and (e)). However, a 20% increase in transmission rate reduction measures from December 1, 2022, would have resulted in a 17.3% (17.4%) reduction in the baseline peak number of daily cases (deaths) with the cases (deaths) attaining a peak about ≈ 7.5 (8.5) months later compared to the baseline scenario (comparing the second peaks of the blue curves with the first peaks of the light green curves in Fig. 10 (b) and (e)). The corresponding percentage reductions in the cumulative cases and deaths by June 30, 2023, will be 3.7% and 1.2%, respectively, (comparing the blue and light green curves in Fig. 11 (b) and (e)). If transmission rate reduction measures were increased by 50% from December 1, 2022, containing the COVID-19 pandemic in the U.S. will be possible (dark green curves in Fig. 10 (b) and (e)). In this case, the number of confirmed daily cases and deaths will go below one by May 21, 2024, and July 5, 2023, respectively, and 6.2% (2.1%) of the baseline cumulative cases (deaths) will be averted by June 30, 2023 (comparing the blue and dark green curves in Fig. 11 (b) and (e)).

Additional simulations were carried out to assess the impact of relaxing and/or reinforcing vaccination and transmission rate reduction measures simultaneously. The results suggest that drastic increases in the daily and cumulative cases and deaths will be registered if the relaxation of these measures is high enough, while the COVID-19 pandemic in the U.S. can be contained if reinforcement of these measures is high enough. In particular, if the number of vaccinated and boosted people per day were reduced by 50% and transmission rate reduction measures were relaxed by 50% starting on December 1, 2022, then a more devastating wave with a daily case (death) peak size that is about ≈ 2.7 (2.5) times the corresponding baseline peak size will be recorded (comparing the second peaks of the blue curves with the first peaks of the gold curves in Fig. 10 (c) and (f)). The corresponding percentage increases in the cumulative cases and deaths by June 30, 2023 will be 13% and 6%, respectively, (comparing the second peaks of the blue curves and the first peaks of the gold curves in Fig. 11 (c) and (f)). On the other hand, if the number of vaccinated and boosted people per day were reduced by 50% and transmission rate reduction measures were reinforced by 50% starting on December 1, 2022, then the number of cases (deaths) for the seventh wave will peak 6 (5) months earlier than the baseline scenario, with a daily case (death) peak size of ≈ 1.8 (1.7) times the corresponding baseline peak size (comparing the second peaks of the blue curves with the first peaks of the light green curves in Fig. 10 (c) and (f)). The corresponding percentage increases in the cumulative cases and deaths by June 30, 2023, will be 9% and 4%, respectively, (comparing the blue and light green curves in Fig. 11 (c) and (f)). Furthermore, if the number of vaccinated and boosted people per day were increased by 50%, while transmission rate reduction measures were relaxed by 50% starting on December 1, 2022, then the number of cases will fall below 100 by October 23, 2023, while the number of deaths will fall below one by August 28, 2023 (comparing the second peaks of the blue curves and the first peaks of the dark green curves in Fig. 10 (c) and (f)). For this scenario, 6% (2%) of the cumulative cases (deaths) will be averted by June 30, 2023 (comparing the blue and dark green curves in Fig. 11 (c) and (f)). Reductions in the daily and cumulative number of cases and deaths will be even more significant, and the time to disease elimination will be reduced if both vaccination and transmission rate reduction measures are reinforced simultaneously. In particular, if the number of people vaccinated per day is increased by 50% and transmission rate reduction measures are also increased by 50% starting on December 1, 2022, then eliminating the COVID-19 pandemic from the U.S. will be possible. Specifically, it will be possible to reduce the number of confirmed new daily cases and deaths below one by January 17 2024 and June 22, 2023, respectively, (dotted purple curves in Fig. 10 (c) and (f)). Also, a 6.2% (2.1%) reduction in the baseline cumulative cases (deaths) will be recorded by June 30, 2023 (comparing the blue and dotted purple curves in Fig. 11 (c) and (f)). It should be mentioned that results on the relaxation of transmission rate reduction measures discussed here are applicable to the case of emergence of a more transmissible VOC.

#### 3.3.5. Assessing the impact of waning natural and vaccine-induced immunity

The model given by Eqs. (2.3) and (2.6) in section 2.5 is simulated using the fixed parameter values in Table S3 and the estimated baseline parameter values in Table S8 of the SI to assess the impact of waning natural immunity on the burden of COVID-19 in the U.S. The results of the simulations (depicted in Figs. 12 (a) and (d) and Fig. 13 (a) and (d)) show that waning of natural immunity has a significant effect on the number of new daily and cumulative COVID-19 cases and deaths in the U.S., in comparison to the baseline scenario. In particular, if natural immunity wanes four times faster (i.e., within 2.25 months in comparison to 9 months for the baseline case), the average number of new daily cases (deaths) at the peak increases by ≈ 101% (≈ 105%), in comparison to the baseline scenario (comparing the peaks of the blue and magenta curves in Fig. 12 (a) and (d)). For this fast-waning scenario, the system settles on the endemic equilibrium faster. However, if natural immunity wanes slowly compared to the baseline scenario, a reduction in the number of cases and deaths at the peak is recorded. In particular, if natural immunity wanes two times slower than the baseline case (i.e., within 18 months in comparison to the 9 months baseline case), the number of new daily cases (deaths) at the peak reduces by ≈ 17% (≈ 17%), in comparison to the baseline scenario (comparing the peaks of the blue and light green curves in Fig. 12 (a) and (d)). The reduction is even more significant if natural immunity wanes faster (comparing the peaks of the blue and dark green curves in Fig. 12 (a) and (d)). Similar changes are obtained for the cumulative cases and deaths (Fig. 13 (a) and (d)). For example, if natural immunity wanes four times faster, a 50% (19%) increase in the number of cumulative cases (deaths), in comparison to the baseline scenario, will be recorded by June 30, 2023 (comparing the blue and magenta curves in Fig. 13 (a) and (d)), while for the case in which natural immunity wanes four times slower than the baseline scenario, an 11% (5%) reduction in the cumulative cases (deaths), in comparison to the baseline case will be recorded by June 30, 2023 (comparing the blue and dark green curves in Fig. 13 (a) and (d)).

**Fig. 12:**
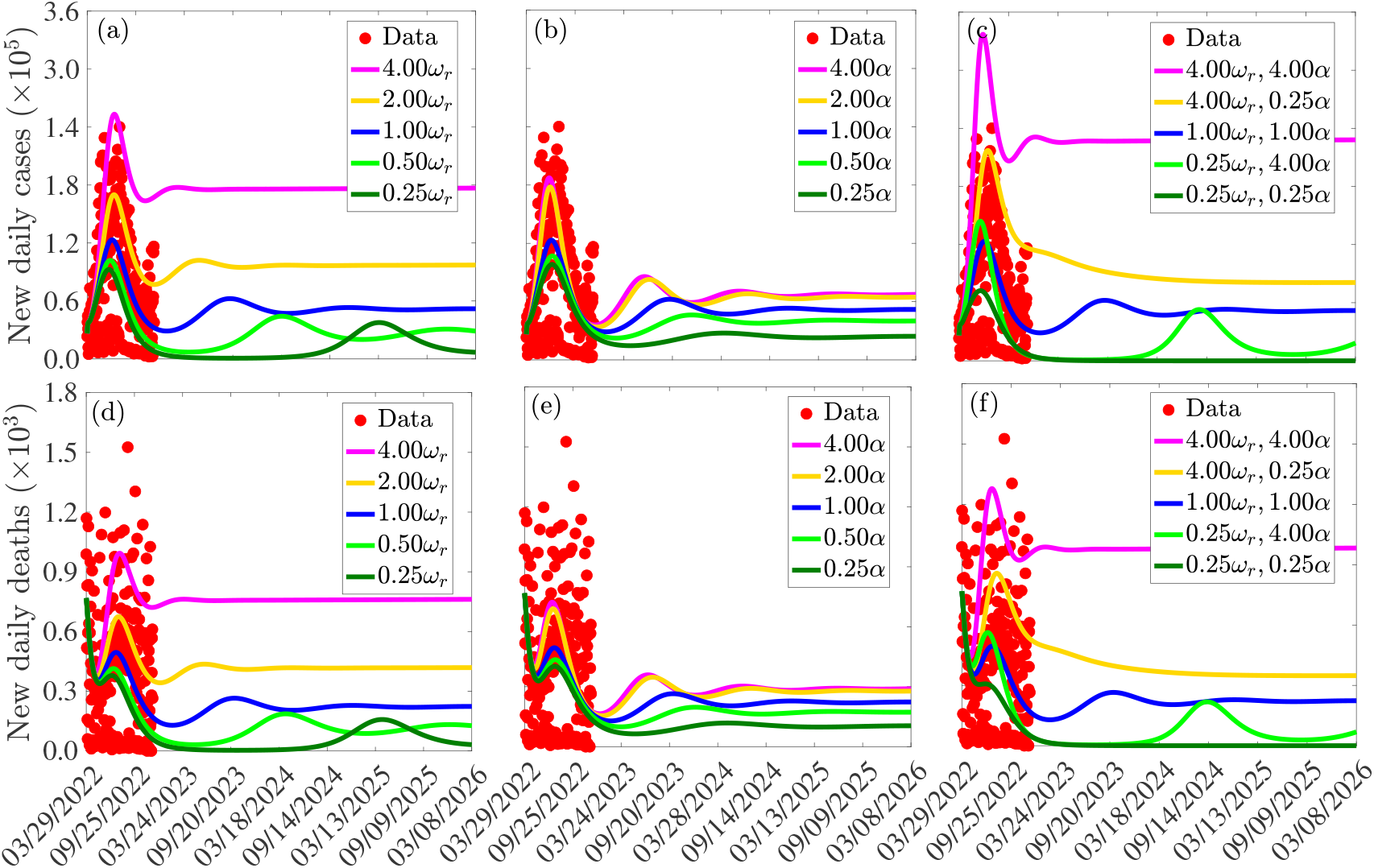
Simulations of Model 4 depicting the impact of waning natural immunity ((a) and (d)), waning vaccine-induced immunity ((b) and (e)), and both waning natural and vaccine-induced immunity ((c) and (f)) on the daily confirmed COVID-19 cases ((a)-(c)) and deaths ((d)-(f)) in the U.S. The parameter (*ω*_*r*_) is the rate at which natural immunity wanes, while *α* is the set of vaccine-induced immunity waning rates. The other parameters used for the simulations are given in Tables S3 and S8 of the SI.

**Fig. 13:**
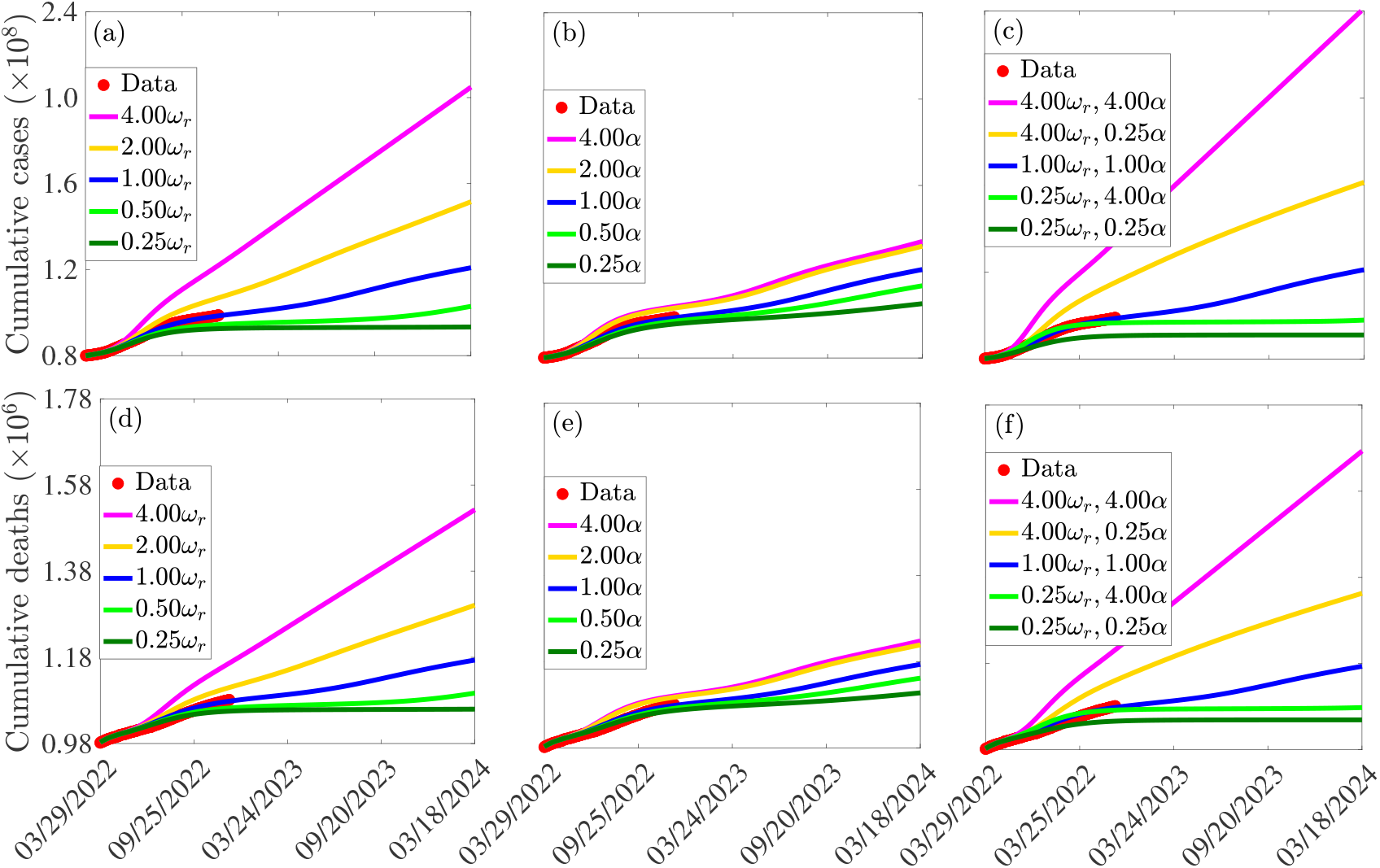
Simulations of the model {(2.3), (2.6)} depicting the impact of waning natural immunity ((a) and (d)), waning vaccine-induced immunity ((b) and (e)), and both waning natural and vaccine-induced immunity ((c) and (f)) on the daily confirmed COVID-19 cases ((a)-(c)) and deaths ((d)-(f)) in the U.S. The parameter (*ω*_*r*_) is the rate at which natural immunity wanes, while *α* is the set of vaccine-induced immunity waning rates. The other parameters used for the simulations are given in Tables S3 and S8 of the SI.

Additionally, Model 4 is simulated using the fixed parameter values in Table S3 and the estimated baseline parameter values in Table S8 of the SI to assess the impact of waning vaccine-induced immunity on the burden of COVID-19 in the U.S. The results obtained and depicted in Figs. 12 (b) and (e) and Fig. 13 (b) and (e) show that waning of vaccine-induced immunity has a lower effect on the number of new daily and cumulative COVID-19 cases and deaths in the U.S. in comparison to waning natural immunity. If vaccine-induced immunity wanes four times faster than the baseline case, the number of new daily cases (deaths) at the peak increases by ≈ 51% (≈ 46%), in comparison to the baseline scenario (comparing the peaks of the blue and magenta curves in Fig. 12 (b) and (e)). For this four times fast waning scenario, the system settles on the endemic equilibrium faster, and a 10% (4%) increase in the baseline number of cumulative cases (deaths) will be recorded by June 30, 2023 (comparing the blue and magenta curves in Fig. 13 (b) and (e)). However, if vaccine-induced immunity wanes four times slower than the baseline scenario, the number of new daily cases (deaths) at the peak reduces by ≈ 20% (≈ 18%), in comparison to the baseline scenario (comparing the peaks of the blue and dark green curves in Fig. 12 (b) and (e)). For this four times slow waning case, a 7% (3%) reduction in the cumulative cases (deaths), in comparison to the baseline case will be recorded by June 30, 2023 (comparing the blue and dark green curves in Fig. 13 (b) and (e)).

Furthermore, the model given by Eqs. (2.3) and (2.6) is simulated using the fixed parameter values in Table S3 and the estimated baseline parameter values in Table S8 of the SI to assess the combined impact of waning natural and vaccine-induced immunity on the burden of COVID-19 in the U.S. The results obtained and depicted in Figs. 12 (c) and (f) and Fig. 13 (c) and (f) show that waning of natural and vaccine-induced immunity has a more significant effect on the number of new daily and cumulative COVID-19 cases and deaths in the U.S. in comparison to waning natural immunity or waning vaccine-induced immunity alone. Specifically, if both natural and vaccine-induced immunity wane four times faster than the respective baseline cases, the number of new daily cases (deaths) at the peak increases by ≈ 174% (≈ 158%), in comparison to the baseline scenario (comparing the peaks of the blue and magenta curves in Fig. 12 (c) and (f)). For this case, the cumulative number of cases (deaths) will increase by ≈ 71% (≈ 28%) in comparison to the baseline scenario by June 30, 2023 (comparing the blue and magenta curves in Fig. 13 (c) and (f)). On the other hand, if natural immunity wanes four times faster and vaccine-induced immunity wanes four times slower, a 76% (74%) increase in the number of new daily cases (deaths) will be recorded at the peak in comparison to the baseline scenario (comparing the peaks of the blue and gold curves in Fig. 12 (c) and (f)), while if natural immunity wanes four times slower and vaccine-induced immunity wanes four times faster than the respective baseline scenarios, a 17% (14%) reduction in the number of new daily cases (deaths) will be recorded at the next peak in comparison to the baseline scenario (comparing the peaks of the blue and light green curves in Fig. 12 (c) and (f)). However, if both natural and vaccine-induced immunity wane four times slower than the respective baseline cases, a 41% (38%) reduction in the number of new daily cases (deaths) will be recorded at the peak in comparison to the baseline scenario (comparing the peaks of the blue and dark green curves in Fig. 12 (c) and (f)), while ≈ 15% (≈ 6%) cumulative cases (deaths) will be averted by June 30, 2023 (comparing the blue and dark green curves in Fig. 13 (c) and (f)).

## 4. Discussion, limitations, and conclusion

### 4.1. Discussion

Over time, different public health interventions have been implemented in the U.S. to reduce the spread of the COVID-19 pandemic. These include quarantine of suspected cases, tracing of contacts of confirmed cases, isolation of confirmed cases, and detection of positive cases through mass testing. In addition to these public health measures, three safe and highly effective vaccines developed, deployed, and administered at warp speed have contributed significantly in curtailing the spread of the virus in the U.S. However, the effectiveness of these vaccines in combating COVID-19 has been threatened by factors such as vaccine hesitancy, the emergence of various variants of concern against which most vaccines designed for the wild-type strain of the virus provide reduced or limited cross-protection, and waning of vaccine-induced immunity over time. This, coupled with the relaxation of NPI mandates in many states across the U.S. since April 20, 2020 made eliminating COVID-19 in the country increasingly difficult. In this study, a library of four mathematical models with increasing complexity is developed and used to study the impact of single- versus double-dose vaccines, the timing of vaccination and boosting, relaxation versus reinforcement of vaccination and transmission reducing NPIs, as well the impact of waning natural and vaccine-induced immunity on the dynamics of COVID-19 in the U.S. The first or basic model accounts for the epidemiological characteristics of the disease and the detection of positive cases. The basic model is then extended to account for vaccination with either a single-dose or a two-dose (mRNA) vaccine and waning vaccine-induced immunity, while subsequent models also account for the boosting of fully vaccinated individuals.

The models are trained with confirmed new daily case and mortality data for the U.S. from the start of the pandemic in the U.S. (i.e., from January 22, 2020) to June 13, 2022, and validated using the portion of the same data set from June 13, 2022 to December 11, 2022, and with cumulative case and mortality data from January 22, 2020 to December 11, 2022. The validation shows an excellent match between the observed daily (cumulative) COVID-19 case and mortality data and the daily (cumulative) cases and deaths from our model. Our model fitting confirms the fact that pre-symptomatic and asymptomatic infectious individuals were responsible for most COVID-19 transmissions in the U.S. This result agrees with previous results reported in [50, 62–64]. Also, the model fitting confirms the fact that transmission of COVID-19 in the U.S. was highest during the main Omicron wave, with the average transmission rate of the Omicron period about twice that of the previous wave and the peak number of cases (death) ≈ 5 (1.4) times that of the previous wave. Additionally, the surge in cases during the omicron wave, as seen in the high control reproduction number, which was ≈ 3.6 times that of the previous wave, demonstrated the epidemiological implication of relaxing the implementation of public health measures, the waning effect of both natural and vaccine-acquired immunity and the emergence of VOC. This result is consistent with those in [23, 101–103]. Also, this finding is similar to that obtained during the first wave, where due to a lack of, and poor adherence to NPIs, the control reproduction number was equally high. In particular, the estimated reproduction numbers for each of the waves from our model are in the range of those reported in [43]. Furthermore, using the fixed and calibrated parameters, it was shown that in the absence of waning vaccine-derived immunity, at least 79% of the population must be fully vaccinated using a combination of the J & J and mRNA vaccines to attain vaccine-induced herd immunity and that this proportion can be as high as 96%, if vaccine-derived immunity wanes over time, even if a small proportion of this fully vaccinated population is vaccinated and boosted twice. Since ≈ 68% of the U.S. population was fully vaccinated, and only about 33% of these fully vaccinated individuals had received at least one booster dose by early December 2022 [19, 31], vaccinating (boosting) a sizeable proportion of the unvaccinated (fully vaccinated but not boosted) population is necessary for achieving vaccine-induced herd immunity in the country.

The extended models with vaccination and booster uptake were simulated using fixed parameters (drawn from the literature) and estimated parameter values to assess the impact of the vaccine type used for vaccination and boosting in the U.S. As expected, the simulation results indicate that using only the more effective mRNA vaccines with an average protective efficacy of 94.55% outperforms using only the J & J vaccine with an average efficacy of 67%. In particular, the study shows that a sizeable number of cases and deaths would have been averted if only mRNA vaccines were used for vaccination and boosting in the U.S. Furthermore, the study shows that ramping up the vaccination rate from when vaccination started would have resulted in a significant reduction in the number of COVID-19 related cases and deaths in the U.S. In particular, if twice the number of people who were vaccinated per day were vaccinated from the onset of vaccination in the U.S., a reduction of almost 30% of the observed cases and deaths would have been recorded on the day that the wave in which vaccine was started peaked and almost one in every two cases and deaths would have been averted on the day that the Delta variant peaked in the U.S. Also, if four times the number of people who received the first booster dose were boosted per day from the onset of boosting in the U.S., a reduction of about one in every four observed cases and about one in every five deaths would have been recorded on the day that the Omicron wave peaked. It should be mentioned that administering booster shots reduced the control reproduction number significantly. In particular, implementing booster shots during the second part of the pandemic wave driven by the Delta variant led to a 44% reduction in the reproduction number (ℛ_*c*3_) of the first portion of the wave (i.e., the period just before the onset of boosting). Also, the effectiveness of the booster doses to mitigate the effect of waning immunity is demonstrated in the wave following the Omicron wave with a significant reduction in the control reproduction number (by ≈ 51%).

Our findings emphasize the fact that early implementation of vaccination and booster administration is critically important in controlling SARS-CoV-2. In other words, late implementation of a vaccination program, especially during the ascendance phase of the Outbreak, would have resulted in a significant increase in the magnitude of the peaks of subsequent waves. Furthermore, simulations of the models show that early and massive implementation of vaccination and booster update policies in the U.S. would have reduced the peak number of cases and deaths significantly. This result agrees with previous studies on the impact of vaccination and NPI timing during the spread of communicable diseases [73, 106, 107]. Unfortunately, the initial administration of vaccination and boosting in the U.S. was low, with about one in three Americans aged 65 and above not yet boosted even by May 11, 2022, although over 90% of individuals within this age bracket were fully vaccinated by May 8, 2022 [82].

The last model (with two booster doses) was simulated to assess the impact of relaxation and reinforcement of vaccination, boosting, and control measures that reduce COVID-19 transmission rates, such as masking up and social distancing in the U.S. The results show that early termination of vaccination and other control measures that reduce the effective transmission rate, or the emergence of another VOC that is more transmissible, can lead to another catastrophic wave of the pandemic depending on the level of relaxation. Also, the study shows that, although both scenarios will generate more cases and deaths compared to the baseline case when the pandemic peaks, relaxing vaccination while reinforcing transmission rate reduction measures by approximately the same percentage will lead to fewer peak numbers of cases and deaths compared to reinforcing vaccination while relaxing transmission rate reduction measures by the same percentage. On the other hand, if vaccination and transmission rate-reducing measures are reinforced simultaneously, elimination of the virus from the U.S. is possible, with the time to elimination determined by the level of reinforcement. Furthermore, the study shows that the waning of natural immunity to COVID-19 has a more significant impact on the number of cases and deaths than the waning of vaccine-derived immunity. This suggests that in addition to boosting vaccine-induced immunity, boosting natural immunity, e.g., through treatment or the use of immune-boosting supplements is important in combating COVID-19 [26, 27].

### 4.2. Limitations (caveats)

The study has some limitations, including assumptions made in building the models, which are important to mention.

- Our models assume homogeneous mixing, in which everyone within the study population has an equal chance of mixing with everyone else within the population.
- It is assumed that individuals in the infected and infectious classes, as well as recovered individuals, are not vaccinated. Also, it is assumed that unvaccinated infectious and vaccinated infectious individuals have the same chance of transmitting the virus to susceptible individuals. Furthermore, it is assumed that when vaccine-induced immunity starts waning, individuals move to another class with a lower vaccine efficacy and that when vaccine-induced immunity wanes completely, individuals progress to the susceptible unvaccinated class. Additionally, it is assumed that individuals who were fully vaccinated with the J & J vaccine pick up the protective efficacy of an mRNA vaccine if they are boosted with an mRNA vaccine.
- Calibration of the unknown parameters of the model was carried out using incidence and mortality data. This is limited in precision due to under-reporting, which is notable for COVID-19 data in the U.S. Also, this is limited compared to fitting a model using wastewater data and could potentially influence the accuracy of our findings.
- Although we acknowledge the impact of age structure on the dynamics of the disease, our focus was on the impact of various vaccines and doses on the entire population. Hence, we did not account for age structure in our study.
- We did not incorporate the possibility of co-infection with both the Omicron and Delta variants.

### 4.3. Conclusion

More than two years since the first case of SARS-CoV-2 infection was reported in China (U.S.) in late December 2019 (January 2020), the goal of NPIs and vaccine policies to contain the disease has not been achieved. Here, we provide quantifiable evidence on the impact of different vaccines and booster shots administered in the U.S. on the incidence of COVID-19 and predict the future effects of the Omicron variant on disease dynamics. Our findings confirm the benefit of early vaccination and booster shots in reducing the pandemic surge. In the absence of vaccination and boosting, subsequent waves will be catastrophic, and the pandemic will remain a major public burden for longer than expected. The administration of vaccines and booster doses reduced the number of cases and deaths significantly. On the other hand, The study shows that although vaccine and booster uptake have been effective in protecting the U.S. populace against COVID-19 infection, as well as severe disease, hospitalization and death when infected, the emergence of an immune evading or more transmissible variant of concern, coupled with waning natural and vaccine-induced immunity, as well as human behavioral changes in response to control measures can result in another COVID-19 wave. Also, the study shows that the response to waning immunity and new variants through booster shots in the country contributed significantly in reducing the number of cases and deaths. Furthermore, the study shows that not relaxing existing control measures prematurely is important, as such relaxation could result in a more devastating outbreak, especially if both vaccination strategies and measures to reduce transmission rates, such as the use of masks, are eased at the same time.

## Data Availability

All data produced in the present study are available upon reasonable request to the authors

## Acknowledgements

CNN acknowledges the support of the Simons Foundation (Award #627346) and the National Science Foundation (Grant Number: DMS #2151870).

