## Supplementary material for "Critical assessment of the impact of vaccines and waning/boosting of immunity on the burden of COVID-19 in the U.S": Online supplementary information

### Online supplementary material

#### 1. Descriptions of the variables and parameters of the model models in Sections 2.2-2.5 of the main paper.

Table S1: Brief descriptions of the variables of the models in Sections 2.2-2.5 of the main paper. Vaccination and boosting is either with the Johnson and Johnson (J & J) vaccine or any of the mRNA (i.e., the Pfizer-BioNTech or Moderna) vaccines.

| Variables | Definition |
|---|---|
| $S_u$ | Unvaccinated susceptible individuals. |
| $S_{ju}$ ($S_{mu}$) | Susceptible individuals who have received only one dose of the J&J (an mRNA) vaccine. |
| $S_{m1}$ | Susceptible individuals who are fully vaccinated with two doses of the same mRNA vaccine |
| $S_{jw1}$ ($S_{mw1}$) | Susceptible individuals who are fully vaccinated with the J&J (an mRNA) vaccine and whose immunity is started to wane. |
| $S_{j2}(S_{m3})$ | Susceptible individuals who are fully vaccinated with the J&J (an mRNA) vaccine, who receive the first J&J (mRNA) booster dose afetr their immunity stats waning. |
| $S_{jm1}$ | Susceptible individuals who are fully vaccinated with the J&J vaccine, who receive an mRNA first booster shot after their immunity has starts waning. |
| $S_{jw2}(S_{mw2})$ | Susceptible individuals who are fully vaccinated and boosted with the first booster dose of the J&J (an mRNA) vaccine, whose immunity has started to wane. |
| $S_{jmw}$ | Susceptible individuals who are fully vaccinated with the J&J vaccine, boosted with the first booster dose the J&J vaccine, who immunity has starts waning. |
| $S_{j3}$ ($S_{m4}$) | Susceptible individuals who are fully vaccinated and boosted twice with the J&J (an mRNA) vaccine. |
| $S_{jm2}$ | Susceptible individuals who are initially fully vaccinated with the J&J (an mRNA) vaccine, but boosted twice an mRNA vaccine. |
| $S_{j2m}$ | Susceptible individuals who are initially fully vaccinated and boosted the first time with the J&J (an mRNA) vaccine, who receive an mRNA vaccine for the second booster. |
| $E$ | Latent individuals; they have contracted the virus but have not started shedding it. |
| $I_p$ | Pre-symptomatic infectious individuals. These are individuals who start shedding SARS-CoV-2 at the end of the latent period but before the end of the incubation period. |
| $I_s$ ($I_a$) | Symptomatic (asymptomatic) infectious individuals; they do (do not) exhibit disease symptoms at the end of the incubation period. |
| $I_c$ | Confirmed or reported cases. |
| $I_h$ | Individuals who are hospitalized because of the disease. |
| $R$ | Individuals who have recovered from infection. |

*Preprint submitted to Elsevier*

Table S2: Brief descriptions of the parameters of the models in Sections 2.2-2.5 of the main paper.

| Parameter | **Definition or description** |
|---|---|
| $\Lambda$ | Human recruitment rate |
| $\mu$ | Human natural death rate |
| $1/\omega_r$ | Average during of natural or infection-induced immunity. |
| $1/\alpha_k$ | Average duration of vaccine-induced immunity for vaccinated or boosted individuals in the $S_k, k \in \{ju, mu, m1, jw1, mw1, j2, jw2, jm1, jmw, mw2, m3, j2m, jm2, mw2, m4\}$ class. |
| $\xi_i(\tilde{\xi}_i)$ | Vaccination rate of susceptible individuals in the $S_i$, $i \in \{u, m1, jw1, m1, mw1, jw2, jmw, mw2\}$ $(i \in \{jw1, jw2\})$ class. |
| $\varepsilon_k$ | Protective efficacy of vaccines for individuals in the $S_k$ class. |
| $\tau_l$ | Covid-19 testing or positivity rate for individuals in the $I_l, l \in \{p, a, s\}$ infectious class. |
| $r(1-r)$ | Proportion of pre-symptomatyic infectious individuals who become symptomatically (asymptomatically) infectious at the end of the incubation period. |
| $\rho_c$ | Hospitalization rate of confirmed infectiousindividuals. |
| $\delta_s(\delta_c)(\delta_h)$ | COVID-19 induced mortality rate for symptomatic (confirmed) (hospitalized) individuals. |
| $1/\sigma_e(1/\sigma_p)$ | Average duration of the latent (Pre-symptomatic) period. |
| $1/\gamma_a(1/\gamma_s)(1/\gamma_c)(1/\gamma_h)$ | Average duration of the infectious period for individuals in the $I_a(I_s)(I_c)(I_h)$ class. |
| $\beta_p(\beta_a)(\beta_s)(\beta_c)(\beta_h)$ | Effective transmission rate for infectious individuals in the $E_p(I_a)(I_s)(I_c)(I_h)$ class. |

### 2. Fixed and estimated baseline parameter values

Table S3: Fixed baseline parameter values of the models. Apart from the vaccine efficacies ($\varepsilon_.$) and the proportion of presymptomatic infectious individuals, who develop COVID-19 symptomatics at the end of the incubation period ($r$), which are dimensionless, all the other parameters have units of *per* day. In this table, $i \in \{m2, m3, jm1, jm2, m4, j2m\}$, $k \in \{j1, j2, j3\}$, and $l \in \{mw1, mw2, jmw\}, \tilde{l} \in \{jw1, jw2\}$.

| **Parameter** | **Value** | **Source** | **Parameter** | **Value** | **Source** |
|---|---|---|---|---|---|
| $\Lambda$ | $1.15 \times 10^4$ | [3] | $\rho_c$ | $1.67 \times 10^{-1}$ | [9] |
| $\mu$ | $3.43 \times 10^{-5}$ | [3] | $\sigma_e, \sigma_p$ | $4.00 \times 10^{-1}$ | [16–18, 20] |
| $r(1-r)$ | $2.0(8.0) \times 10^{-1}$ | [4, 5] | $\varepsilon_{m1}$ | $5.20 \times 10^{-1}$ | [13] |
| $\omega_r$ | $3.70 \times 10^{-3}$ | [6–8] | $\varepsilon_i$ | $9.46 \times 10^{-1}$ | [14] |
| $\alpha_k$ | $5.56 \times 10^{-3}$ | [6–8] | $\varepsilon_k$ | $6.70 \times 10^{-1}$ | [12] |
| $\alpha_l$ | $1.67 \times 10^{-2}$ | [26] | $\varepsilon_l$ | $7.00 \times 10^{-1}$ | [12] |
| $\alpha_{\tilde{l}}$ | $1.11 \times 10^{-2}$ | [26] | $\varepsilon_{\tilde{l}}$ | $5.00 \times 10^{-1}$ | [12] |

Table S4: Estimated parameter (Par) values and the associated 95% Confidence intervals (CI) of the basic model (i.e., the model no vaccination and boosting) in Section 2.2 of the main paper during the first wave (January 22 to June 1, 2020) and second wave (June 2 to September 14, 2020) of the COVID-19 in the U.S.

(a) First wave (January 22 to June 1, 2020).

| Par | Value | 95% Confidence interval |
|---|---|---|
| $\beta_p$ | $7.4723 \times 10^{-1}$ | $[4.7429 \times 10^{-1}, 8.6174 \times 10^{-1}]$ |
| $\beta_a$ | $5.0479 \times 10^{-1}$ | $[1.2557 \times 10^{-1}, 8.0603 \times 10^{-1}]$ |
| $\beta_s$ | $1.7654 \times 10^{-2}$ | $[2.2871 \times 10^{-6}, 4.3665 \times 10^{-1}]$ |
| $\beta_c$ | $8.8270 \times 10^{-4}$ | $[1.1436 \times 10^{-6}, 2.1832 \times 10^{-2}]$ |
| $\beta_h$ | $1.7654 \times 10^{-4}$ | $[2.2871 \times 10^{-7}, 4.3665 \times 10^{-3}]$ |
| $\tau_p$ | $2.6356 \times 10^{-4}$ | $[3.3516 \times 10^{-6}, 2.8917 \times 10^{-4}]$ |
| $\tau_a$ | $1.0046 \times 10^{-5}$ | $[8.5728 \times 10^{-6}, 2.3169 \times 10^{-4}]$ |
| $\tau_s$ | $5.1581 \times 10^{-4}$ | $[4.9818 \times 10^{-4}, 5.8846 \times 10^{-4}]$ |
| $\delta_s$ | $1.8292 \times 10^{-5}$ | $[6.0482 \times 10^{-6}, 4.1756 \times 10^{-5}]$ |
| $\delta_c$ | $5.9899 \times 10^{-3}$ | $[2.1109 \times 10^{-6}, 3.4328 \times 10^{-2}]$ |
| $\delta_h$ | $6.1436 \times 10^{-3}$ | $[6.9564 \times 10^{-6}, 2.4206 \times 10^{-2}]$ |
| $\gamma_a$ | $2.8477 \times 10^{-1}$ | $[1.4246 \times 10^{-1}, 4.0000 \times 10^{-1}]$ |
| $\gamma_s$ | $8.9074 \times 10^{-3}$ | $[8.3012 \times 10^{-3}, 8.1243 \times 10^{-2}]$ |
| $\gamma_c$ | $1.0704 \times 10^{-1}$ | $[1.8488 \times 10^{-6}, 3.9604 \times 10^{-1}]$ |
| $\gamma_h$ | $3.8143 \times 10^{-1}$ | $[1.1126 \times 10^{-4}, 4.1940 \times 10^{-1}]$ |
| $\mathscr{R}_{c1}$ | 3.66 | $[1.89, 4.83]$ |

(b) Second wave (June 2 to September 14, 2020).

| Par | Value | 95% Confidence interval |
|---|---|---|
| $\beta_p$ | $3.5879 \times 10^{-1}$ | $[3.6236 \times 10^{-2}, 6.1083 \times 10^{-1}]$ |
| $\beta_a$ | $7.0237 \times 10^{-2}$ | $[2.1346 \times 10^{-3}, 1.8045 \times 10^{-1}]$ |
| $\beta_s$ | $3.2660 \times 10^{-2}$ | $[1.0216 \times 10^{-3}, 1.6196 \times 10^{-1}]$ |
| $\beta_c$ | $1.6330 \times 10^{-3}$ | $[5.1080 \times 10^{-5}, 8.0980 \times 10^{-3}]$ |
| $\beta_h$ | $3.6601 \times 10^{-4}$ | $[1.0216 \times 10^{-5}, 1.6196 \times 10^{-3}]$ |
| $\tau_p$ | $3.7201 \times 10^{-3}$ | $[3.0158 \times 10^{-5}, 7.4460 \times 10^{-3}]$ |
| $\tau_a$ | $3.2191 \times 10^{-4}$ | $[1.0457 \times 10^{-5}, 4.3871 \times 10^{-4}]$ |
| $\tau_s$ | $5.4991 \times 10^{-4}$ | $[4.4538 \times 10^{-5}, 7.8313 \times 10^{-4}]$ |
| $\delta_s$ | $2.8053 \times 10^{-5}$ | $[2.2843 \times 10^{-6}, 1.6917 \times 10^{-4}]$ |
| $\delta_c$ | $8.4844 \times 10^{-4}$ | $[2.4092 \times 10^{-6}, 1.2514 \times 10^{-3}]$ |
| $\delta_h$ | $1.8931 \times 10^{-3}$ | $[3.4425 \times 10^{-6}, 8.5385 \times 10^{-3}]$ |
| $\gamma_a$ | $1.9647 \times 10^{-2}$ | $[1.1321 \times 10^{-3}, 4.2286 \times 10^{-2}]$ |
| $\gamma_s$ | $1.0983 \times 10^{-1}$ | $[2.1145 \times 10^{-3}, 3.0443 \times 10^{-1}]$ |
| $\gamma_c$ | $3.6232 \times 10^{-1}$ | $[4.3887 \times 10^{-3}, 3.1394 \times 10^{-1}]$ |
| $\gamma_h$ | $4.2416 \times 10^{-2}$ | $[3.2893 \times 10^{-3}, 3.0894 \times 10^{-1}]$ |
| $\mathscr{R}_{c1}$ | 3.73 | $[1.63, 4.92]$ |

Table S5: Estimated parameter (Par) values and the associated 95% Confidence intervals for the basic model (i.e., the model with no vaccination and boosting) in Section 2.2 of the main paper during the first part of the third of the COVID-19 pandemic in the U.S. (i.e., from September 15 to December 18, 2020).

| Par | Value | 95% Confidence interval |
|---|---|---|
| $\beta_p$ | $1.5640 \times 10^{-1}$ | $[4.5599 \times 10^{-2}, 2.9521 \times 10^{-1}]$ |
| $\beta_a$ | $5.2391 \times 10^{-2}$ | $[1.4953 \times 10^{-2}, 9.2373 \times 10^{-2}]$ |
| $\beta_s$ | $1.0014 \times 10^{-3}$ | $[2.9534 \times 10^{-4}, 8.9545 \times 10^{-1}]$ |
| $\beta_c$ | $5.0070 \times 10^{-5}$ | $[1.4767 \times 10^{-5}, 4.4773 \times 10^{-2}]$ |
| $\beta_h$ | $1.0014 \times 10^{-5}$ | $[2.9534 \times 10^{-6}, 8.9545 \times 10^{-3}]$ |
| $\tau_p$ | $1.0239 \times 10^{-2}$ | $[1.8203 \times 10^{-4}, 1.9830 \times 10^{-2}]$ |
| $\tau_a$ | $5.7933 \times 10^{-4}$ | $[1.3891 \times 10^{-5}, 2.5070 \times 10^{-3}]$ |
| $\tau_s$ | $3.0410 \times 10^{-2}$ | $[1.8505 \times 10^{-4}, 7.9210 \times 10^{-2}]$ |
| $\delta_s$ | $3.2689 \times 10^{-5}$ | $[2.1924 \times 10^{-6}, 3.3091 \times 10^{-3}]$ |
| $\delta_c$ | $3.0260 \times 10^{-3}$ | $[2.4120 \times 10^{-5}, 6.1973 \times 10^{-3}]$ |
| $\delta_h$ | $2.4571 \times 10^{-3}$ | $[4.6300 \times 10^{-4}, 3.2677 \times 10^{-2}]$ |
| $\gamma_a$ | $8.3522 \times 10^{-3}$ | $[4.6104 \times 10^{-3}, 3.4114 \times 10^{-2}]$ |
| $\gamma_s$ | $1.8844 \times 10^{-1}$ | $[4.2812 \times 10^{-2}, 4.1921 \times 10^{-1}]$ |
| $\gamma_c$ | $2.9343 \times 10^{-1}$ | $[2.1556 \times 10^{-3}, 3.9810 \times 10^{-1}]$ |
| $\gamma_h$ | $1.6697 \times 10^{-1}$ | $[4.1930 \times 10^{-3}, 3.8175 \times 10^{-1}]$ |
| $\mathscr{R}_{c1}$ | 4.94 | $[2.68, 5.38]$ |

Table S6: Estimated parameter (Par) values and the associated 95% Confidence intervals for the vaccination model with no booster uptake in Section 2.3 of the main paper during the second part of the third wave (i.e., from December 19, 2020 to July 4, 2021) and part of the fourth wave (i.e., from July 5, to September 24, 2021) of the COVID-19 in the U.S.

(a) Segment from December 19, 2020 to July 3, 2021.

| Par | Value | 95% Confidence interval |
|---|---|---|
| $\beta_p$ | $7.1498 \times 10^{-1}$ | $[5.8230 \times 10^{-1}, 9.2184 \times 10^{-1}]$ |
| $\beta_a$ | $3.0139 \times 10^{-2}$ | $[1.1431 \times 10^{-2}, 2.7198 \times 10^{-2}]$ |
| $\beta_s$ | $1.1130 \times 10^{-2}$ | $[1.0000 \times 10^{-2}, 2.9595 \times 10^{-1}]$ |
| $\beta_c$ | $5.5650 \times 10^{-4}$ | $[5.0000 \times 10^{-4}, 1.4798 \times 10^{-1}]$ |
| $\beta_h$ | $1.1130 \times 10^{-4}$ | $[1.0000 \times 10^{-4}, 2.9595 \times 10^{-2}]$ |
| $\tau_p$ | $6.4222 \times 10^{-6}$ | $[3.2675 \times 10^{-9}, 8.8929 \times 10^{-3}]$ |
| $\tau_a$ | $8.4925 \times 10^{-4}$ | $[6.6252 \times 10^{-4}, 1.1032 \times 10^{-3}]$ |
| $\tau_s$ | $5.4873 \times 10^{-2}$ | $[4.5939 \times 10^{-2}, 6.8613 \times 10^{-2}]$ |
| $\delta_s$ | $4.2648 \times 10^{-4}$ | $[0.0000 \times 10^{-0}, 2.4385 \times 10^{-3}]$ |
| $\delta_c$ | $2.1490 \times 10^{-7}$ | $[0.0000 \times 10^{-0}, 5.1872 \times 10^{-3}]$ |
| $\delta_h$ | $1.1141 \times 10^{-3}$ | $[7.0422 \times 10^{-9}, 5.3773 \times 10^{-3}]$ |
| $\gamma_a$ | $1.4840 \times 10^{-2}$ | $[1.1661 \times 10^{-2}, 1.7221 \times 10^{-2}]$ |
| $\gamma_s$ | $7.8336 \times 10^{-2}$ | $[5.3226 \times 10^{-2}, 1.2157 \times 10^{-1}]$ |
| $\gamma_c$ | $7.2077 \times 10^{-2}$ | $[4.0510 \times 10^{-9}, 3.3332 \times 10^{-1}]$ |
| $\gamma_h$ | $7.2681 \times 10^{-2}$ | $[6.6667 \times 10^{-2}, 1.9998 \times 10^{-1}]$ |
| $\xi_{ju}$ | $1.6781 \times 10^{-3}$ | $[1.5468 \times 10^{-8}, 3.4475 \times 10^{-2}]$ |
| $\xi_{mu}$ | $5.2991 \times 10^{-2}$ | $[3.2288 \times 10^{-2}, 7.0939 \times 10^{-2}]$ |
| $\xi_{m1}$ | $1.19205 \times 10^{-1}$ | $[7.3075 \times 10^{-2}, 2.3466 \times 10^{-1}]$ |
| $\mathscr{R}_{c2}$ | 0.94 | $[0.69, 1.18]$ |

(b) Segment from July 5, to September 24, 2021.

| Par | Value | 95% Confidence interval |
|---|---|---|
| $\beta_p$ | $7.2882 \times 10^{-1}$ | $[2.6891 \times 10^{-2}, 9.8861 \times 10^{-1}]$ |
| $\beta_a$ | $9.8526 \times 10^{-2}$ | $[1.3483 \times 10^{-2}, 7.8594 \times 10^{-1}]$ |
| $\beta_s$ | $6.3700 \times 10^{-1}$ | $[4.1393 \times 10^{-3}, 9.6529 \times 10^{-1}]$ |
| $\beta_c$ | $3.1850 \times 10^{-2}$ | $[2.0697 \times 10^{-4}, 4.8265 \times 10^{-2}]$ |
| $\beta_h$ | $6.3700 \times 10^{-3}$ | $[4.1393 \times 10^{-5}, 9.6529 \times 10^{-3}]$ |
| $\tau_p$ | $2.2576 \times 10^{-3}$ | $[2.5542 \times 10^{-4}, 1.0205 \times 10^{-2}]$ |
| $\tau_a$ | $1.0383 \times 10^{-3}$ | $[2.6060 \times 10^{-5}, 4.6898 \times 10^{-3}]$ |
| $\tau_s$ | $1.6314 \times 10^{-2}$ | $[4.5174 \times 10^{-5}, 2.5731 \times 10^{-2}]$ |
| $\delta_s$ | $6.3076 \times 10^{-7}$ | $[2.3810 \times 10^{-5}, 1.6695 \times 10^{-3}]$ |
| $\delta_c$ | $1.2731 \times 10^{-4}$ | $[2.2391 \times 10^{-5}, 9.4059 \times 10^{-3}]$ |
| $\delta_h$ | $9.0347 \times 10^{-4}$ | $[3.0265 \times 10^{-5}, 9.8477 \times 10^{-3}]$ |
| $\gamma_a$ | $4.2740 \times 10^{-2}$ | $[9.6311 \times 10^{-3}, 3.8392 \times 10^{-1}]$ |
| $\gamma_s$ | $5.7216 \times 10^{-2}$ | $[1.1680 \times 10^{-2}, 1.6839 \times 10^{-1}]$ |
| $\gamma_c$ | $4.4351 \times 10^{-3}$ | $[3.5239 \times 10^{-4}, 2.3294 \times 10^{-1}]$ |
| $\gamma_h$ | $6.6667 \times 10^{-2}$ | $[2.1693 \times 10^{-2}, 2.1985 \times 10^{-1}]$ |
| $\xi_{ju}$ | $3.6222 \times 10^{-3}$ | $[4.2615 \times 10^{-5}, 8.2527 \times 10^{-3}]$ |
| $\xi_{mu}$ | $2.5120 \times 10^{-2}$ | $[1.8921 \times 10^{-4}, 8.4563 \times 10^{-2}]$ |
| $\xi_{m1}$ | $3.7897 \times 10^{-2}$ | $[3.7148 \times 10^{-4}, 6.1847 \times 10^{-2}]$ |
| $\mathscr{R}_{c2}$ | 1.78 | $[1.22, 2.10]$ |

Table S7: Estimated parameter (Par) values and the associated 95% Confidence intervals for the vaccination model with first booster uptake in Section 2.4 of the main paper during the periods from September 25 to December 2, 2021 and December 3, 2021 to March 28, 2022.

(a) Segment from September 25 to December 2, 2021.

| Par | Value | 95% Confidence interval |
|---|---|---|
| $\beta_p$ | $4.4803 \times 10^{-1}$ | $[1.7932 \times 10^{-1}, 7.6173 \times 10^{-1}]$ |
| $\beta_a$ | $3.5106 \times 10^{-1}$ | $[1.2278 \times 10^{-2}, 4.8721 \times 10^{-1}]$ |
| $\beta_s$ | $1.7435 \times 10^{-1}$ | $[1.5144 \times 10^{-3}, 2.9716 \times 10^{-1}]$ |
| $\beta_c$ | $8.7175 \times 10^{-3}$ | $[7.5720 \times 10^{-5}, 1.4858 \times 10^{-2}]$ |
| $\beta_h$ | $1.7435 \times 10^{-3}$ | $[1.5144 \times 10^{-5}, 2.9716 \times 10^{-3}]$ |
| $\tau_p$ | $1.6167 \times 10^{-2}$ | $[9.0552 \times 10^{-3}, 2.3576 \times 10^{-2}]$ |
| $\tau_a$ | $7.8211 \times 10^{-3}$ | $[3.5100 \times 10^{-5}, 1.6855 \times 10^{-2}]$ |
| $\tau_s$ | $1.1229 \times 10^{-2}$ | $[2.7290 \times 10^{-5}, 5.6805 \times 10^{-2}]$ |
| $\delta_s$ | $6.4985 \times 10^{-5}$ | $[1.7324 \times 10^{-5}, 6.3827 \times 10^{-3}]$ |
| $\delta_c$ | $1.0131 \times 10^{-4}$ | $[1.6002 \times 10^{-5}, 3.8758 \times 10^{-3}]$ |
| $\delta_h$ | $5.8131 \times 10^{-4}$ | $[1.2380 \times 10^{-5}, 2.1640 \times 10^{-3}]$ |
| $\gamma_a$ | $2.84306 \times 10^{-1}$ | $[3.0235 \times 10^{-2}, 1.2483 \times 10^{-1}]$ |
| $\gamma_s$ | $4.9520 \times 10^{-2}$ | $[1.4286 \times 10^{-2}, 1.4286 \times 10^{-1}]$ |
| $\gamma_c$ | $1.6667 \times 10^{-2}$ | $[1.2039 \times 10^{-2}, 1.2494 \times 10^{-1}]$ |
| $\gamma_h$ | $6.6668 \times 10^{-2}$ | $[2.1394 \times 10^{-2}, 2.4821 \times 10^{-1}]$ |
| $\xi_{ju}$ | $7.3059 \times 10^{-3}$ | $[1.0231 \times 10^{-5}, 9.6417 \times 10^{-2}]$ |
| $\xi_{mu}$ | $1.3240 \times 10^{-2}$ | $[1.0178 \times 10^{-5}, 9.7001 \times 10^{-2}]$ |
| $\xi_{m1}$ | $3.8181 \times 10^{-1}$ | $[3.7906 \times 10^{-2}, 4.8420 \times 10^{-1}]$ |
| $\xi_{mw1}$ | $1.0017 \times 10^{-3}$ | $[1.5310 \times 10^{-5}, 7.6949 \times 10^{-2}]$ |
| $\xi_{jw1}$ | $8.7104 \times 10^{-6}$ | $[1.3313 \times 10^{-6}, 6.6912 \times 10^{-3}]$ |
| $\tilde{\xi}_{jw1}$ | $1.0017 \times 10^{-3}$ | $[1.5310 \times 10^{-5}, 7.6949 \times 10^{-2}]$ |
| $\mathscr{R}_{c3}$ | 0.99 | $[0.77, 1.08]$ |

(b) Segment December 3, 2021 to March 28, 2022.

| Par | Value | 95% Confidence interval |
|---|---|---|
| $\beta_p$ | $1.6667 \times 10^{0}$ | $[7.6535 \times 10^{-1}, 2.7932 \times 10^{0}]$ |
| $\beta_a$ | $3.2671 \times 10^{-3}$ | $[1.2769 \times 10^{-3}, 8.1375 \times 10^{-1}]$ |
| $\beta_s$ | $1.2007 \times 10^{-3}$ | $[1.2280 \times 10^{-3}, 5.2614 \times 10^{-1}]$ |
| $\beta_c$ | $6.0040 \times 10^{-5}$ | $[2.2666 \times 10^{-5}, 2.6307 \times 10^{-2}]$ |
| $\beta_h$ | $1.2007 \times 10^{-5}$ | $[6.1400 \times 10^{-5}, 5.2614 \times 10^{-3}]$ |
| $\tau_p$ | $5.0548 \times 10^{-3}$ | $[2.2666 \times 10^{-7}, 6.4592 \times 10^{-3}]$ |
| $\tau_a$ | $1.0220 \times 10^{-6}$ | $[1.0022 \times 10^{-6}, 8.8279 \times 10^{-3}]$ |
| $\tau_s$ | $2.0385 \times 10^{-1}$ | $[1.4817 \times 10^{-3}, 4.9589 \times 10^{-2}]$ |
| $\delta_s$ | $1.0391 \times 10^{-6}$ | $[4.2120 \times 10^{-6}, 1.0075 \times 10^{-3}]$ |
| $\delta_c$ | $1.6084 \times 10^{-4}$ | $[2.2358 \times 10^{-6}, 5.6929 \times 10^{-3}]$ |
| $\delta_h$ | $1.8539 \times 10^{-4}$ | $[1.7010 \times 10^{-6}, 7.7322 \times 10^{-3}]$ |
| $\gamma_a$ | $1.4286 \times 10^{-2}$ | $[5.1429 \times 10^{-3}, 1.6898 \times 10^{-1}]$ |
| $\gamma_s$ | $1.4286 \times 10^{-2}$ | $[6.4286 \times 10^{-3}, 1.4286 \times 10^{-1}]$ |
| $\gamma_c$ | $1.4286 \times 10^{-2}$ | $[4.2857 \times 10^{-3}, 1.2500 \times 10^{-1}]$ |
| $\gamma_h$ | $3.3338 \times 10^{-2}$ | $[3.3357 \times 10^{-3}, 1.4286 \times 10^{-1}]$ |
| $\xi_{ju}$ | $1.3251 \times 10^{-6}$ | $[1.2352 \times 10^{-6}, 6.2800 \times 10^{-5}]$ |
| $\xi_{mu}$ | $9.1077 \times 10^{-4}$ | $[1.7232 \times 10^{-6}, 4.2256 \times 10^{-3}]$ |
| $\xi_{m1}$ | $5.2847 \times 10^{-2}$ | $[1.2857 \times 10^{-4}, 8.3081 \times 10^{-2}]$ |
| $\xi_{mw1}$ | $3.2795 \times 10^{-2}$ | $[1.7372 \times 10^{-4}, 4.3528 \times 10^{-2}]$ |
| $\xi_{jw1}$ | $2.8518 \times 10^{-3}$ | $[1.5106 \times 10^{-5}, 3.7850 \times 10^{-3}]$ |
| $\tilde{\xi}_{jw1}$ | $3.2795 \times 10^{-2}$ | $[1.7372 \times 10^{-4}, 4.3528 \times 10^{-2}]$ |
| $\mathscr{R}_{c3}$ | 3.59 | $[2.14, 5.76]$ |

Table S8: Estimated parameter (Par) values and the associated 95% Confidence intervals for the vaccination model with first booster uptake in Section 2.5 of the main paper during the period from March 29 to June 13, 2022.

| Par | Value | 95% Confidence interval | Par | Value | 95% Confidence interval |
|---|---|---|---|---|---|
| $\beta_p$ | $1.8252\times10^{-1}$ | $[1.3122\times10^{-2}, 1.8479\times10^{0}]$ | $\gamma_c$ | $1.6684\times10^{-2}$ | $[1.4524\times10^{-2}, 1.2495\times10^{-1}]$ |
| $\beta_a$ | $5.6188\times10^{-1}$ | $[5.6578\times10^{-2}, 9.3142\times10^{-1}]$ | $\gamma_h$ | $4.0002\times10^{-2}$ | $[1.8622\times10^{-4}, 1.4286\times10^{-1}]$ |
| $\beta_s$ | $2.5365\times10^{-1}$ | $[1.6527\times10^{-3}, 5.4382\times10^{-1}]$ | $\xi_{ju}$ | $9.2895\times10^{-2}$ | $[4.2318\times10^{-6}, 9.8505\times10^{-2}]$ |
| $\beta_c$ | $1.2013\times10^{-2}$ | $[8.2635\times10^{-5}, 2.7191\times10^{-2}]$ | $\xi_{mu}$ | $9.6408\times10^{-2}$ | $[5.3869\times10^{-3}, 9.9854\times10^{-2}]$ |
| $\beta_h$ | $2.5365\times10^{-3}$ | $[1.6527\times10^{-5}, 5.4382\times10^{-3}]$ | $\xi_{m1}$ | $1.8895\times10^{-2}$ | $[4.0421\times10^{-3}, 5.6094\times10^{-2}]$ |
| $\tau_p$ | $4.0401\times10^{-3}$ | $[2.1240\times10^{-6}, 8.5038\times10^{-3}]$ | $\xi_{mw1}$ | $1.6899\times10^{-3}$ | $[1.2366\times10^{-4}, 3.4051\times10^{-2}]$ |
| $\tau_a$ | $1.1202\times10^{-3}$ | $[5.9816\times10^{-6}, 1.2259\times10^{-2}]$ | $\xi_{jw1}$ | $1.4695\times10^{-4}$ | $[1.0753\times10^{-5}, 2.9610\times10^{-3}]$ |
| $\tau_s$ | $3.1450\times10^{-2}$ | $[5.3728\times10^{-3}, 6.9301\times10^{-2}]$ | $\tilde{\xi}_{jw1}$ | $1.6899\times10^{-3}$ | $[1.2366\times10^{-4}, 3.4051\times10^{-2}]$ |
| $\delta_s$ | $3.4862\times10^{-5}$ | $[1.3234\times10^{-8}, 1.8537\times10^{-3}]$ | $\xi_{mw2}$ | $1.2013\times10^{-3}$ | $[1.0208\times10^{-5}, 7.9189\times10^{-2}]$ |
| $\delta_c$ | $1.9476\times10^{-5}$ | $[2.0369\times10^{-8}, 1.7349\times10^{-3}]$ | $\xi_{jmw}$ | $1.2013\times10^{-3}$ | $[1.0208\times10^{-5}, 7.9189\times10^{-2}]$ |
| $\delta_h$ | $1.4388\times10^{-4}$ | $[4.1066\times10^{-8}, 2.0685\times10^{-3}]$ | $\xi_{jw2}$ | $1.0446\times10^{-3}$ | $[8.8765\times10^{-7}, 6.8860\times10^{-3}]$ |
| $\gamma_a$ | $1.0377\times10^{-1}$ | $[3.5931\times10^{-2}, 2.3190\times10^{-1}]$ | $\tilde{\xi}_{jw2}$ | $1.2013\times10^{-3}$ | $[1.0208\times10^{-5}, 7.9189\times10^{-2}]$ |
| $\gamma_s$ | $2.5548\times10^{-2}$ | $[1.1757\times10^{-2}, 6.6191\times10^{-2}]$ | $\mathscr{R}_{c4}$ | 1.76 | $[0.85, 2.25]$ |

### 3. Initial conditions

It should be mentioned that from the second wave onward, some of the initial conditions were estimated, while others were the end points of the previous wave. Only the estimated initial conditions are stated here.

#### 3.1. *Initial conditions for Model 1*

The initial conditions for the first wave are $S_u(0) = 336218628$, $E(0) = 10$, $I_p(0) = 10$, $I_a(0) = 1$, $I_s(0) = 10$, $I_c(0) = 1$, $I_h(0) = 0$, $R(0) = 0$, while the estimated initial conditions for the second wave are $S_u(0) = 295346686$, $I_a(0) = 1828780$, $I_c(0) = 22545$, $I_h(0) = 145000$, $R(0) = 20000$. The initial conditions for the portion of the third wave before the start of vaccination are: $S_u(0) = 329036214$, $E(0) = 2778164$, $I_p(0) = 2831272$, $I_a(0) = 1000000$, $I_s(0) = 12028$, $I_c(0) = 37418$, $I_h(0) = 292467$, $R(0) = 30000$.

#### 3.2. *Initial conditions for Model 2*

The estimated initial conditions for the period in which vaccination started are $S_{ju}(0) = 0$, $S_{mu}(0) = 948695$, $S_{m1}(0) = 0$, $S_{jw1}(0) = 0$, $S_{mw1}(0) = 0$, $E(0) = 1914615$, $I_p(0) = 74866$, $I_a(0) = 73867311$, $I_c(0) = 246520$, $I_h(0) = 1178565$, $R(0) = 7009633$, while the estimated initial conditions for the period of the Delta variant wave before the start of boosting are $I_c(0) = 24652$, $I_h(0) = 438652$, $R(0) = 13297843$

#### 3.3. *Initial conditions for Model 3*

The initial conditions for the second part of the Delta wave, i.e., the period during which boosting started are: $S_{j2}(0) = 128564$, $S_{jw2}(0) = 0$, $S_{m3}(0) = 2956970$, $S_{mw2}(0) = 0$, $S_{jm1}(0) = 128564$, $S_{jmw}(0) = 0$, $I_p(0) = 6450841$, $I_a(0) = 1237474$, $I_s(0) = 52739$, $I_c(0) = 122519$, $I_h(0) = 3262581$, while the initial conditions for the Omicron wave are: $S_{ju}(0) = 22068935$, $S_{mu}(0) = 1375929$, $S_{m1}(0) = 79254287$, $S_{jw1}(0) = 74660040$, $S_{mw1}(0) = 20863255$, $S_{j2}(0) = 2374927$, $S_{jw2}(0) = 48700$, $S_{m3}(0) = 1607049$, $S_{mw2}(0) = 81140$, $S_{jm1}(0) = 1607049$, $S_{jmw}(0) = 9738795$, $E(0) = 429866$, $I_p(0) = 427914$, $I_a(0) = 587613$, $I_s(0) = 191874$, $I_c(0) = 125793$, $I_h(0) = 3482156$.

#### 3.4. *Initial conditions for Model 4*

The initial conditions for the second booster period are: $S_{ju}(0) = 17107851$, $S_{mu}(0) = 41272838$, $S_{m1}(0) = 42979145$, $S_{jw1}(0) = 65383476$, $S_{mw1}(0) = 14660615$, $S_{j2}(0) = 27196223$, $S_{jw2}(0) = 92182792$, $S_{m3}(0) = 13503009$, $S_{mw2}(0) = 85240864$, $S_{jm1}(0) = 11484000$, $S_{jmw}(0) = 15111253$, $S_{j3}(0) = 11484$, $S_{jm2}(0) = 1511125$, $S_{m4}(0) = 3022251$, $S_{j2m}(0) = 1511125$, $E(0) = 302225$, $I_p(0) = 551115$, $I_a(0) = 136423$, $I_s(0) = 1.27181$, $I_c(0) = 226977$, $I_h(0) = 5276800$, $R_h(0) = 75000000$,

### 4. Supplementary figures

#### *4.1. Assessing the impact of the type of vaccine used for vaccination and boosting*

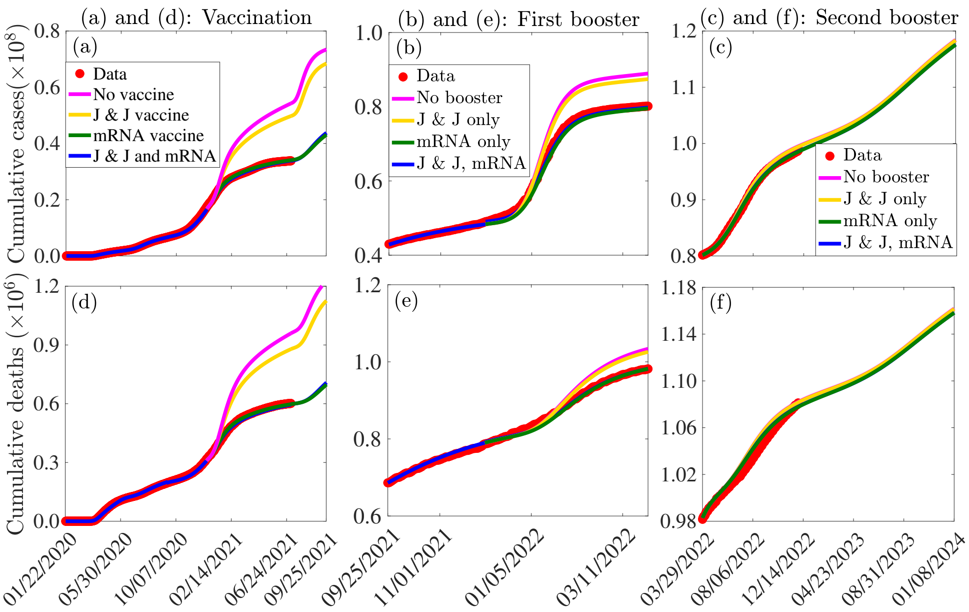


**Fig.** S1: Simulations of Models 1-4 depicting the impact on the cumulative cases ((a)-(c)) and deaths ((d)-(f)) of using only the J & J (magenta curve), Pfizer-BioNTech or Moderna vaccines (dark green curves), or both the J & J and Pfizer-BioNTech or Moderna vaccines (blue curve) for vaccinating unvaccinated individuals ((a) and (d)), first booster uptake for fully vaccinated individuals ((b) and (e)), and for boosting individuals, who have already received first booster shots ((c) and (f)). The other parameters used for the simulations are presented in Tables S3-S8.

#### *4.2. Assessing the impact of vaccination and booster timing*

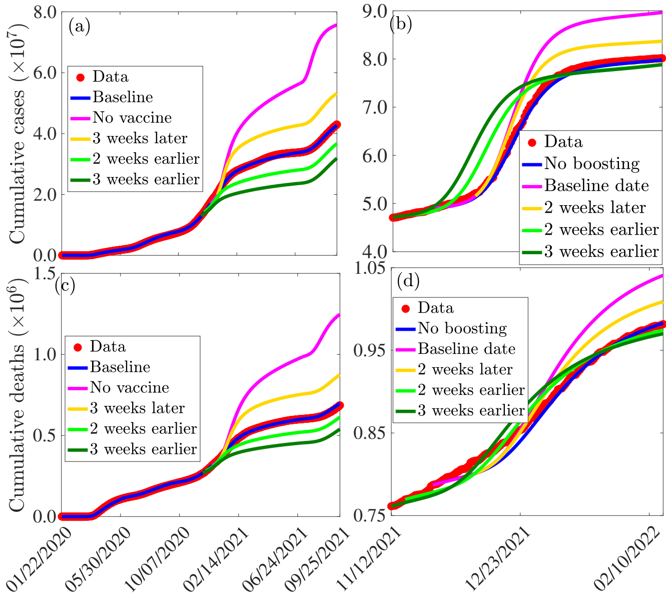


**Fig.** S2: Simulations of Models 1-4 depicting the impact of administration timing of vaccination ((a) and (c)) and first booster update ((b) and (d)) on the cumulative number of COVID-19 cases ((a)-(b)) and the cumulative number of COVID-19 deaths ((c)-(d)) in the U.S. The parameters used for the simulations are presented in Tables S3-S8 in the SI.

#### *4.3. Assessing the impact of vaccine and booster uptake*

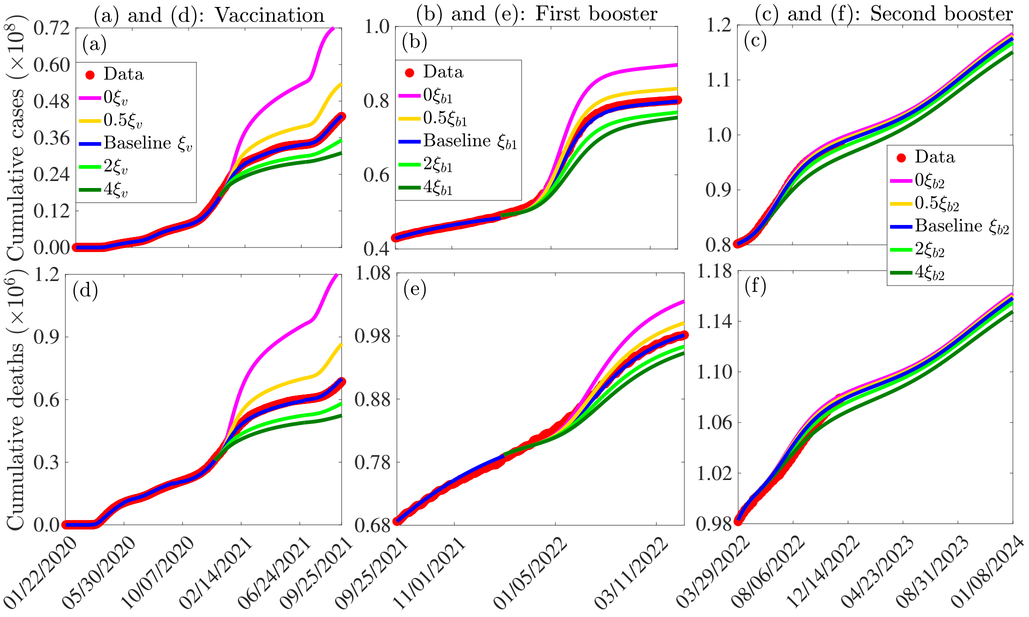


**Fig.** S3: Simulations of Models 2, 3 and 4 to assess the impact on vaccination ((a) and (d)), first booster uptake ((b) and (e)), and second booster uptake ((c) and (f)), on the cumulative confirmed cases ((a) and (c)) and deaths ((e) and (f)) The vaccination rate ($\xi_v$) is given by $\xi_v = \{\xi_{ju}, \xi_{mu}, \xi_{m1}\}$, the rate at which first booster shots are administered ($\xi_{b1}$) is given by $\xi_{b1} = \{\xi_{jw1}, \tilde{\xi}_{jw1}, \xi_{mw1}\}$, and the rate at which second booster shots are administered ($\xi_{b2}$) is given by $\xi_{b2} = \{\xi_2, \tilde{\xi}_{jw2}, \xi_{jmw}, \xi_{mw2}\}$. The other parameter values used for the simulations are given in Tables S3-S8.

#### *4.4. Assessing the impact of relaxing or reinforcing control measures implemented in the U.S.*

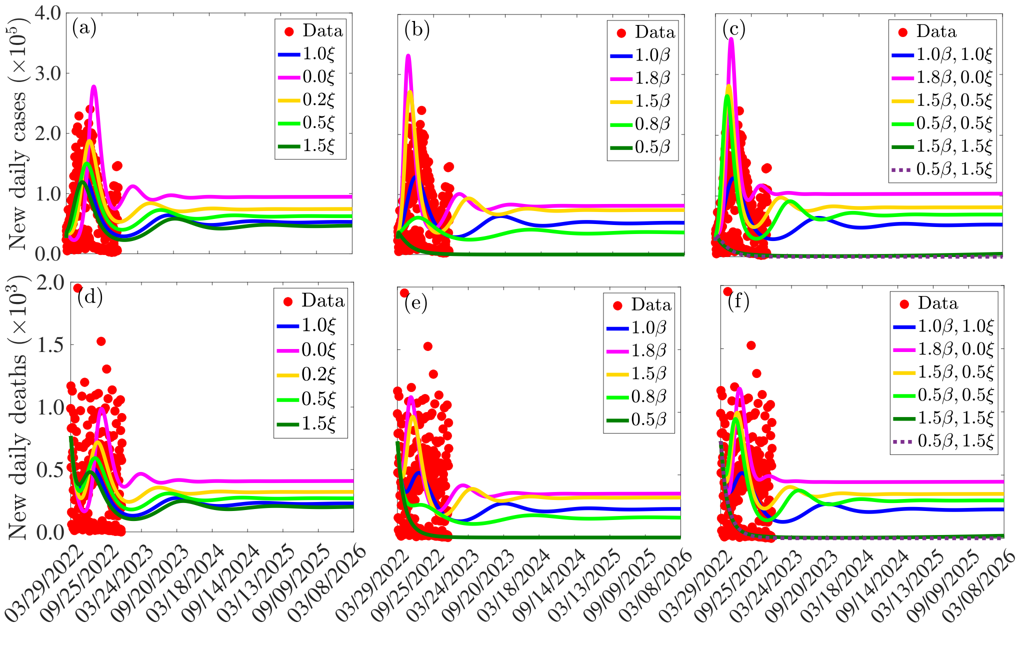


**Fig.** S4: Simulations of Model 4 depicting the impact of relaxation/reinforcement in vaccination ((a) and (d)), transmission reducing control measures ((b) and (e)), and both vaccination and transmission reducing measures ((c) and (f)) on the daily confirmed COVID-19 cases ((a)-(c)) and deaths ((d)-(f)) in the U.S. The vector $\xi$ entries are the baseline vaccination rates associated with mRNA and the J & J vaccines, while $\beta = \{\beta_p, \beta_a, \beta_c, \beta_c, \beta_h\}$. Hypothetical scenario in which relaxation and/or reinforcement of measures started on March 29, 2022. The other parameters used for the simulations are given in Tables S3 and S8 in the SI.

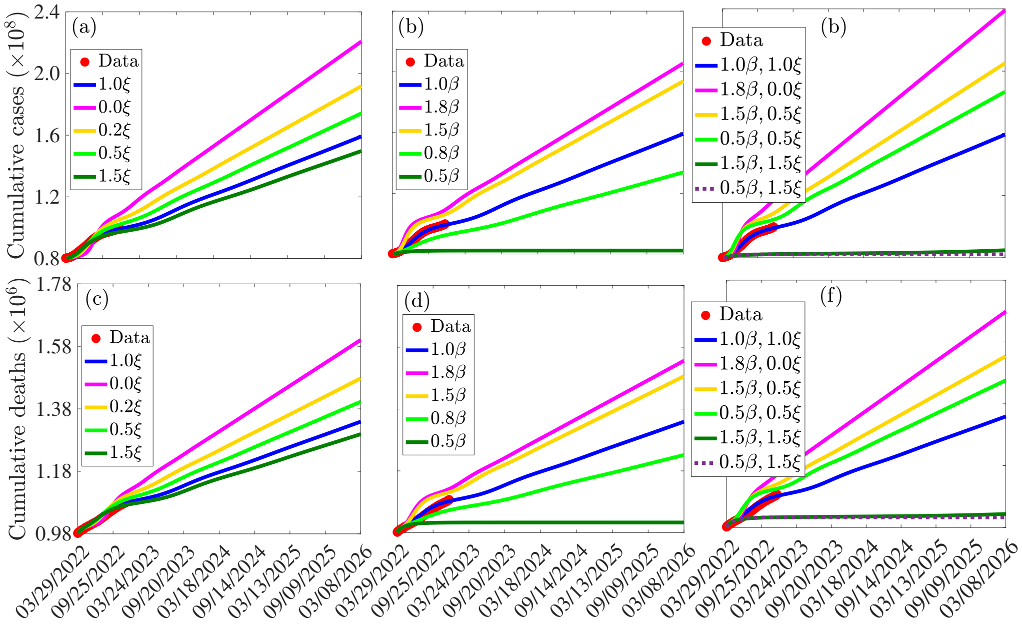


**Fig.** S5: Simulations of Model 4 depicting the impact of relaxation/reinforcement in vaccination ((a) and (d)), transmission rate reducing measures ((b) and (e)), and both vaccination and transmission reducing measures ((c) and (f)) on the daily confirmed COVID-19 cases ((a)-(c)) and deaths ((d)-(f)) in the U.S. The entries of the vector $\xi$ are the baseline vaccination rates associated with mRNA and the J & J vaccines, while $\beta = \{\beta_p, \beta_a, \beta_c, \beta_c, \beta_h\}$. Hypothetical scenario in which relaxation and/or reinforcement of measures started on March 29, 2022. The other parameters used for the simulations are given in Tables S3 and S8 in the SI.
